# Swab-Seq: A high-throughput platform for massively scaled up SARS-CoV-2 testing

**DOI:** 10.1101/2020.08.04.20167874

**Authors:** Joshua S. Bloom, Laila Sathe, Chetan Munugala, Eric M. Jones, Molly Gasperini, Nathan B. Lubock, Fauna Yarza, Erin M. Thompson, Kyle M. Kovary, Jimin Park, Dawn Marquette, Stephania Kay, Mark Lucas, TreQuan Love, A. Sina Booeshaghi, Oliver F. Brandenberg, Longhua Guo, James Boocock, Myles Hochman, Scott W. Simpkins, Isabella Lin, Nathan LaPierre, Duke Hong, Yi Zhang, Gabriel Oland, Bianca Judy Choe, Sukantha Chandrasekaran, Evann E. Hilt, Manish J. Butte, Robert Damoiseaux, Clifford Kravit, Aaron R. Cooper, Yi Yin, Lior Pachter, Omai B. Garner, Jonathan Flint, Eleazar Eskin, Chongyuan Luo, Sriram Kosuri, Leonid Kruglyak, Valerie A. Arboleda

## Abstract

The rapid spread of severe acute respiratory syndrome coronavirus 2 (SARS-CoV-2) is due to the high rates of transmission by individuals who are asymptomatic at the time of transmission^1, 2^. Frequent, widespread testing of the asymptomatic population for SARS-CoV-2 is essential to suppress viral transmission. Despite increases in testing capacity, multiple challenges remain in deploying traditional reverse transcription and quantitative PCR (RT-qPCR) tests at the scale required for population screening of asymptomatic individuals. We have developed SwabSeq, a high-throughput testing platform for SARS-CoV-2 that uses next-generation sequencing as a readout. SwabSeq employs sample-specific molecular barcodes to enable thousands of samples to be combined and simultaneously analyzed for the presence or absence of SARS-CoV-2 in a single run. Importantly, SwabSeq incorporates an *in vitro* RNA standard that mimics the viral amplicon, but can be distinguished by sequencing. This standard allows for end-point rather than quantitative PCR, improves quantitation, reduces requirements for automation and sample-to-sample normalization, enables purification-free detection, and gives better ability to call true negatives. After setting up SwabSeq in a high-complexity CLIA laboratory, we performed more than 80,000 tests for COVID-19 in less than two months, confirming in a real world setting that SwabSeq inexpensively delivers highly sensitive and specific results at scale, with a turn-around of less than 24 hours. Our clinical laboratory uses SwabSeq to test both nasal and saliva samples without RNA extraction, while maintaining analytical sensitivity comparable to or better than traditional RT-qPCR tests. Moving forward, SwabSeq can rapidly scale up testing to mitigate devastating spread of novel pathogens.

## INTRODUCTION

Public health strategies remain essential tools for controlling the spread of severe acute respiratory syndrome coronavirus 2 (SARS-CoV-2), the cause of COVID-19. In contrast to SARS-CoV-1, for which infectivity is associated with symptoms^3, 4^, infectivity of SARS-CoV-2 is high during the asymptomatic/presymptomatic phase^5, 6^. As a consequence, containing transmission based solely on symptoms is impossible, which makes molecular screening for SARS-CoV-2 essential for pandemic control.

During the surges of infection in the US, the rise in cases has frequently overwhelmed the capacity of the quantitative RT-PCR (qRT-PCR) tests that make up the majority of FDA-authorized tests for COVID-19. Delays in obtaining test results, which are due to capacity constraints rather than assay times^8^, render testing ineffective for the public health aims of preventing viral transmission and suppressing local outbreaks. Even where expanded testing capacity exists, the ∼$100 price of tests (current Medicare reimbursement rates^9^) prohibits their widespread adoption by large employers and schools on a regular basis for effective viral suppression^10, 11^. Frequent, low-cost population testing, combined with contact tracing and isolation of infected individuals, would help to halt the spread of COVID-19 and reopen society^12, 13^. Here we describe SwabSeq, a SARS-CoV-2 testing platform that leverages next-generation sequencing to massively scale up testing capacity^14, 15^.

SwabSeq improves on one-step reverse transcription and polymerase chain reaction (RT-PCR) approaches in several key areas. Like other sequencing approaches, SwabSeq utilizes molecular barcodes that are embedded in the RT-PCR primers to uniquely label each sample and allow for simultaneous sequencing of hundreds to thousands of samples in a single run (LampSeq^16^, Illumina CovidSeq^17^, DxSeq^18^). SwabSeq uses very short reads, reducing sequencing times so that results can be returned in less than 24 hours.

To deliver robust and reliable results at scale, SwabSeq adds to every sample a synthetic *in vitro* RNA standard that has a sequence nearly identical to the target in the virus genome, but is easily distinguished by sequencing. SARS-CoV-2 detection is based on the ratio of the counts of true viral sequencing reads to those from the *in vitro* viral standard. Since every sample contains the synthetic RNA, SwabSeq controls for failure of amplification: samples with no SARS-CoV-2 detected are those in which only *in vitro* viral standard reads are observed, while those without viral or *in vitro* viral standard reads are inconclusive.

The RNA control confers a number of additional advantages on the SwabSeq assay. Since we are only interested in the ratio of real virus to *in vitro* standard, the PCR can be run to the endpoint, where all primers are consumed, rather than for a set number of cycles. By driving the reaction to endpoint, we overcome the presence of varying amounts of RT and PCR inhibitors and effectively force each sample to have similar amounts of final product. Using *in vitro* standard RNA with end-point PCR has two important consequences. First, we can pool reaction products after PCR because the yield of product per sample is mostly determined by the primer concentration and not by the sample concentration. Second, we can directly process extraction-free samples. Inhibitors of RT and PCR present in mucosal tissue or saliva should affect both the virus and the *in vitro* standard equally. Endpoint PCR overcomes the effect of inhibition, while keeping the ratio of reads between the two RNA species approximately constant, and so avoids the need for extraction.

Here we show that SwabSeq has extremely high sensitivity and specificity for the detection of viral RNA in purified samples. We also demonstrate a low limit of detection in extraction-free lysates from mid-nasal swabs and saliva. Finally, we describe our real-world use of SwabSeq to perform more than 80,000 tests for COVID-19 in less than two months in a high-complexity CLIA laboratory. These results demonstrate the potential of SwabSeq to be used for SARS-CoV-2 testing on an unprecedented scale, offering a potential solution to the need for population-wide testing to stem the pandemic.

## RESULTS

SwabSeq is a simple and scalable protocol, consisting of 5 steps (**Figure 1A**): (1) sample collection, (2) reverse transcription and PCR using primers that contain unique molecular indices at the i7 and i5 positions (**Figure 1B, Figure S1**) as well as *in vitro* standards, (3) simple pooling (no normalization) and cleanup of the uniquely barcoded samples for library preparation, (4) sequencing of the pooled library, and (5) computational assignment of barcoded sequencing reads to each sample for counting and viral detection.

**Figure 1.**
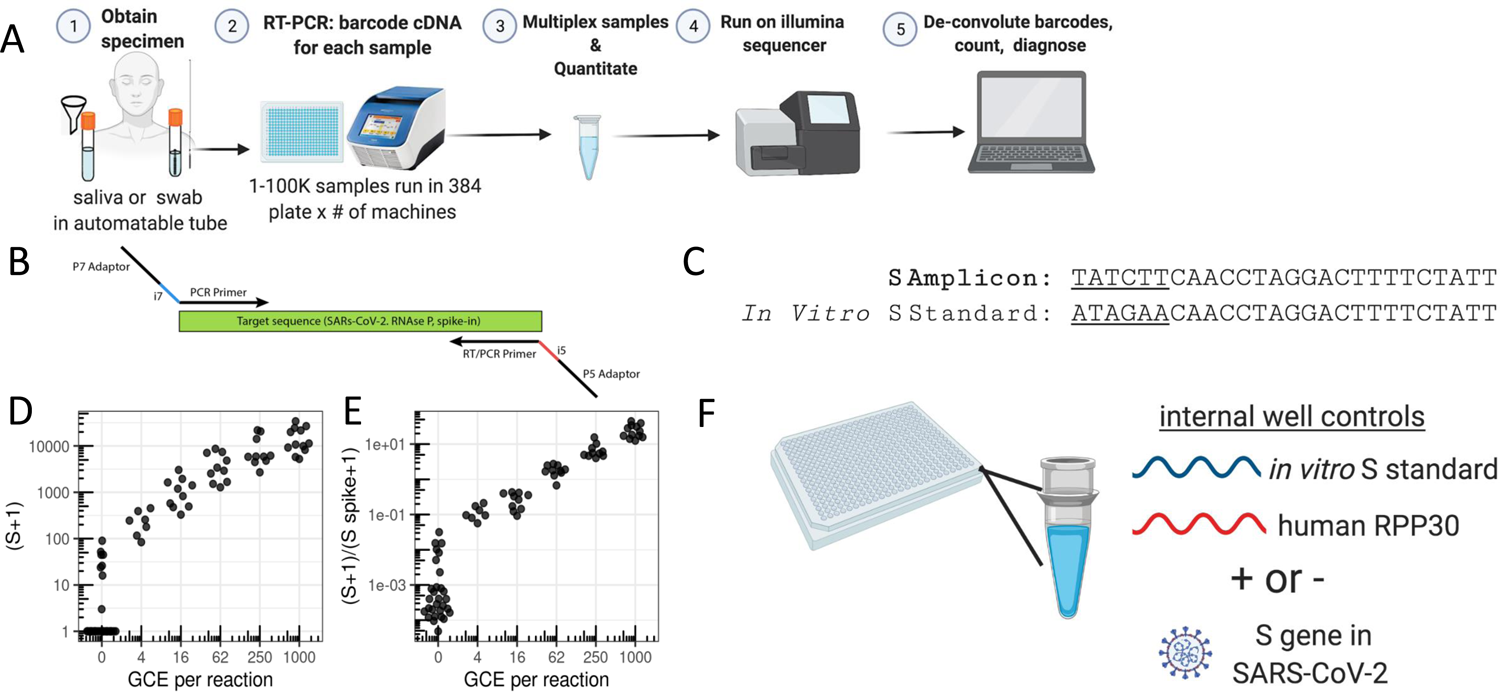
SwabSeq Diagnostic Testing Platform for COVID19. A) The workflow for SwabSeq is a five step process that takes approximately 12 hours from start to finish. B) In each well, we perform RT-PCR on clinical samples. Each well has two sets of indexed primers that generate cDNA and amplicons for SARS-CoV-2 S gene and the human RPP30 gene. Each primer is synthesized with the P5 and P7 adaptors for Illumina sequencing, a unique i7 and i5 molecular barcodes, and the unique primer pair. Importantly, every well has a synthetic *in vitro* S standard that is key to allowing the method to work at scale. C) The *in vitro* S standard (abbreviated as S-Spike) differs from the virus S gene by 6 base pairs that are complemented (underlined). (D) Read count at various viral concentrations (E) Ratiometric normalization allow for in-well normalization for each amplicon (F) Every well has two internal well controls for amplification, the *in vitro* S standard and the human RPP30. The RPP30 amplicon serves as a control for specimen collection. The *in vitro* S standard is critical to SwabSeq’s ability to distinguish true negatives.

Our assay consists of two primer sets that amplify two genes: the S gene of SARS-CoV-2 and the human *Ribonuclease P/MRP Subunit P30* (*RPP30*). We include a synthetic *in vitro* RNA standard that is identical to the viral sequence targeted for amplification, except for the most upstream 6 bp (**Figure 1C**), which allows us to distinguish sequencing reads corresponding to the *in vitro* standard from those corresponding to the target sequence. The primers amplify both the viral and the synthetic sequences with equal efficiency (**Figure S2**). We have also designed a second RNA standard for RPP30 with a similar design. The ratio of the number of native reads to the number of *in vitro* standard reads provides a more accurate and quantitative measure of the number of viral genomes in the sample than native read counts alone (**Figure 1D**,**E**). The *in vitro* standard also allows us to retain linearity over a large range of viral input despite the use of endpoint PCR (**Figure S3**). With this approach, the final amount of DNA in each well is largely defined by the total primer concentration rather than by the viral input—negative samples have higher amounts of *in vitro* S standard (abbreviated as S-spike) and lower/zero amounts of viral reads, and positive samples have lower amounts of *in vitro* S standard and higher amounts of viral reads (**Figure S3**). In addition to viral S, we reverse-transcribe and amplify a human housekeeping gene to control for specimen quality, as in traditional qPCR assays (**Figure 1F**).

We also assign i5 and i7 sample barcodes that are designed to be at least several base-pair edits away from one another, allowing for demultiplexing even in the face of sequencing errors. After RT-PCR, samples are combined at equal volumes, purified, and used to generate one sequencing library. We have used the Illumina MiSeq, MIniSeq, and the Illumina NextSeq 550 to sequence these libraries (**Figure S4**). We minimize instrument sequencing time by sequencing only the minimum required 26 base pairs (**Methods**). Each read is classified as deriving from native or *in vitro* standard S, or RPP30, and assigned to a sample based on the associated index sequences (barcodes). To maximize specificity and avoid false-positive signals arising from incorrect classification or assignment, conservative edit-distance thresholds are used for this matching operation (**Methods** and **Supplemental Results**). A sequencing read is discarded if it does not match one of the expected sequences. Counts for native and *in vitro* standard S and RPP30 reads are obtained for each sample and used for downstream analyses (**Methods**).

We have estimated that approximately 5,000 reads per well are sufficient to detect the presence or absence of viral RNA in a sample (**Methods**). This translates to at least 1,500 samples per run on a MiSeq or MiniSeq, 20,000 samples per run on a NextSeq 550, and up to 150,000 samples per run on a NovaSeq S2 flow cell. Computational analysis takes only minutes per run^19^. The MiniSeq newly released high output rapid flow cell has a turn around time of 2.5 hours, which significantly improves our throughput and turn-around time. Our optimized protocols allow for a single person to process 6-384 well plates in a single hour; which is equivalent to 2304 samples an hour. This process cloned over multiple liquid handlers can rapidly scale to 10,000 samples per hour with a staff of 6 people. In our current operations, we routinely run up to 1536 samples per flow cell on a MiniSeq.

We have optimized the SwabSeq protocol by identifying and eliminating multiple sources of noise (**Supplemental Results**) to create a streamlined and scalable protocol for SARS-CoV-2 testing.

### Validation of SwabSeq as a diagnostic platform

We first validated SwabSeq on purified RNA nasopharyngeal (NP) samples that were previously tested by the UCLA Clinical Microbiology Laboratory with a standard RT-qPCR assay (ThermoFisher Taqpath COVID19 Combo Kit). To determine our analytical limit of detection, we diluted inactivated virus with pooled, remnant clinical NP swab specimens. The remnant samples were all confirmed to be negative for SARS-CoV-2. In these remnant samples, we performed a serial, 2-fold dilution of heat-inactivated SARS-CoV-2 (ATCC® VR-1986HK), from 8,000 to 125 genome copy equivalents (GCE) per mL. We detected SARS-CoV-2 in 34/34 samples down to 250 GCE per mL, and in 28/34 samples down to 125 GCE per mL (**Figure 2A**). These results established that SwabSeq is highly sensitive, with an analytical limit of detection (LOD) of 250 GCE per mL for purified RNA from nasal swabs. This limit of detection is lower than those of many currently FDA authorized and highly sensitive RT-qPCR assays for SARS-CoV-2.

**Figure 2.**
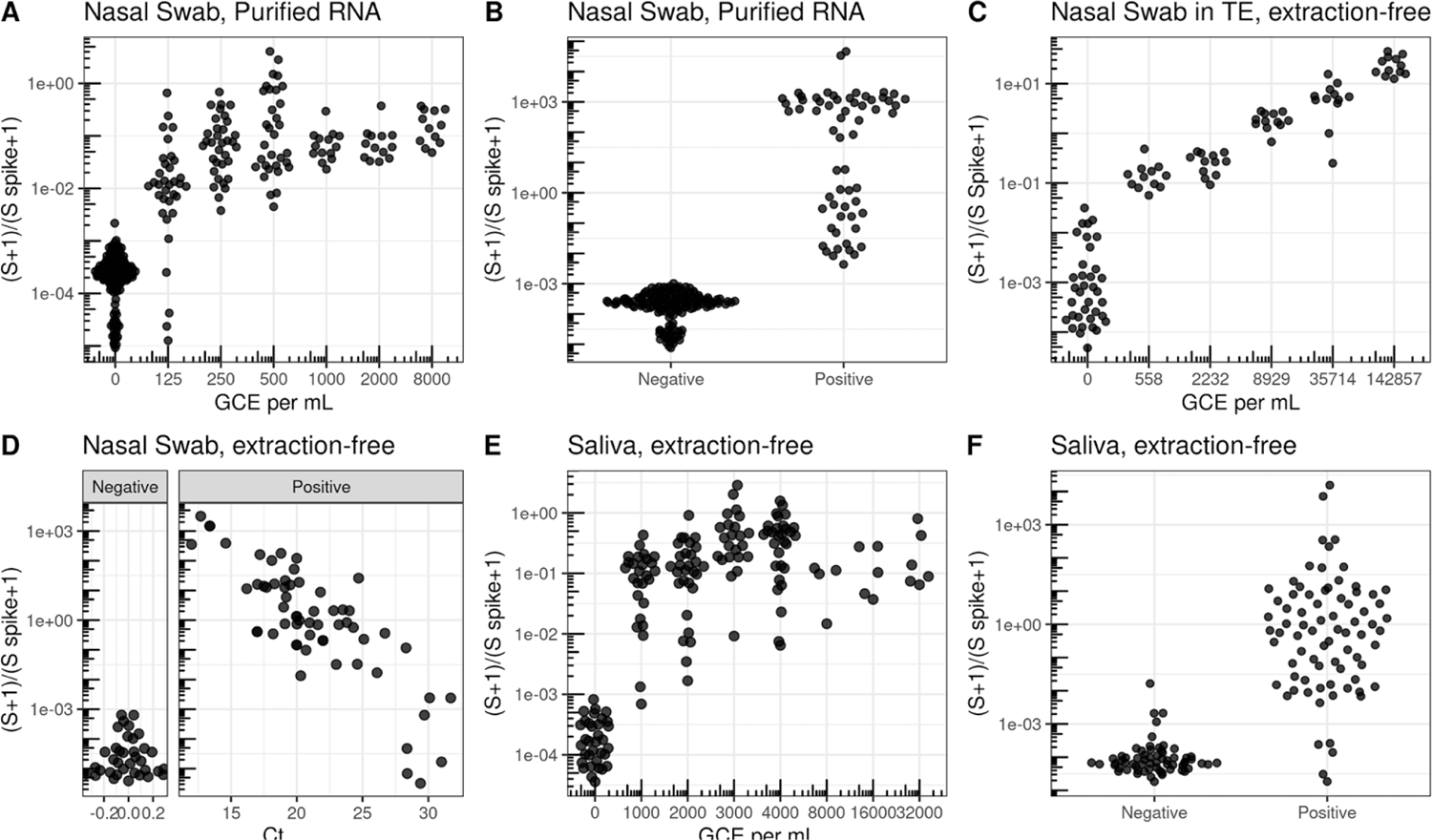
Validation in clinical specimens demonstrate a limit of detection equivalent to sensitive RT-qPCR reactions. A) Limit of Detection in nasal swab samples with no SARS-CoV2 were pooled and ATCC inactivated virus was added at different concentrations. Nasal Swab sample was RNA purified and using SwabSeq showed a limit of detection of 250 genome copy equivalents (GCE) per mL. B) RNA-purified clinical nasal swab specimens obtained through the UCLA Health Clinical Microbiology Laboratory were tested based on clinical protocols using FDA authorized platforms and then also tested using SwabSeq. This represents a subset of the total purified RNA samples used in our validation. We show 100% agreement with samples that tested positive for SARS-CoV-2 (n=63) and negative for SARS-CoV-2 (n=159). C) We also tested RNA purified samples from extraction-free nasopharyngeal swab and showed a limit of detection of 558 GCE/mL. D) Relationship between Ct from RT-qPCR targeting the S gene (x-axis) and SwabSeq ratio for extract-free swabs into normal saline or Tris-EDTA (y-axis) for patient samples classified as testing positive or negative for SARS-CoV-2 by the UCLA Clinical Microbiology Laboratory. Samples with no virus detected were assigned a Ct of 0 for this visualization. E) Extraction-free processing of saliva specimens show a limit or detection down to 1000 GCE per mL. F) Extraction-free processing of saliva clinical specimens using swabseq (y-axis) compared to classification of SARS-CoV-2 status from RNA-purified clinical nasal swab specimens for matched samples (x-axis).

SwabSeq detects the SARS-CoV-2 genome with high clinical sensitivity and specificity. We retested 380 RNA-purified nasopharyngeal samples from the UCLA Clinical Microbiology Laboratory in which SARS-CoV-2 positive (n=94) and negative (n=286). We observed 99.2% agreement with RT-qPCR results for all samples (**Figure 2B** and **S5D**) with 100% concordance in samples that were positive for SARS-CoV-2. We sequenced these samples on either a MiSeq, MiniSeq or on a NextSeq550 (**Figure S5**), with 100% concordance between the different sequencing instruments.

One of the major bottlenecks in scaling up RT-qPCR diagnostic tests is the RNA purification step. RNA extraction is challenging to automate, making it a time consuming and labor intensive step, and supply chains have not been able to keep up with the demand for the necessary reagents during the course of the pandemic. Thus, we explored the ability of SwabSeq to detect SARS-CoV-2 directly from a variety of extraction-free sample types. The CDC recommends several types of media for nasal swab collection: viral transport medium (VTM)^20^, Amies transport medium^21^, and normal saline^21^. A main technical challenge arises from RT or PCR inhibition by ingredients in these collection buffers. We found that diluting specimens with water overcame the RT and PCR inhibition and allowed us to detect viral RNA in contrived and positive clinical patient samples, with higher limits of detections between 4000 and 6000 GCE/mL (**Figure S6**). We also tested nasal swabs that were collected directly into Tris-EDTA (TE) buffer, diluted 1:1 with water. This approach yielded a limit of detection of 560 GCE/mL (**Figure 2C**). We next performed a comparison between traditional purified RT-qPCR protocols and our extraction free-protocol for nasopharyngeal samples collected into either normal saline (n=128) or TE (n=170). We showed 96.0% overall agreement, 92.0% positive percent agreement and 97.6% negative percent agreement for all samples and a high correlation (r^2^=0.68) between SwabSeq signal and RT-qPCR, and high reproducibility at the lower end of our LOD (**Figure 2D, Figures S7-S9**).

We also tested extraction-free saliva protocols in which saliva is collected directly into a matrix tube using a funnel-like collection device (**Figure 1A**). The main technical challenges in demonstrating the detection of virus in saliva samples have been preventing degradation of the inactivated SARS-CoV-2 virus that is added to saliva and ensuring accurate pipetting of this heterogeneous and viscous sample type. We found that heating the saliva samples to 95℃ for 30 minutes^22^ reduced PCR inhibition and improved detection of the S amplicon (**Figure S10**). After heating, we diluted samples at a 1:1 volume with 2x TBE with 1% Tween-20^22^. Using this method, we obtained a LoD of 1000 GCE/mL (**Figure 2E**). In a study performed in the UCLA Emergency Department, we collected paired saliva and nasopharyngeal swab samples in patients with COVID19-like symptoms and compared our extraction-free, saliva SwabSeq protocol and purified nasopharyngeal swab samples run with high-sensitivity RT-qPCR in the UCLA Clinical Microbiology lab. We showed 95.3% overall agreement, 90.2% positive percent agreement, and 96.3% negative percent agreement across 537 samples. (**Figure S11**).

### Deployment of SwabSeq for large-scale asymptomatic screening

After obtaining a CLIA license on October 30, 2020, we carried out a pilot phase of clinical testing starting in November 2020, during which we tested 8,199 samples over 6 weeks (**Figure 3A**). Following the success of this pilot phase, we began large-scale testing at the start of 2021 to support asymptomatic screening of health care workers at a major academic medical center as well as students and staff at local colleges and universities. To date, we have returned results for over 80,000 tests, at a scale of ∼10,000 tests per week, with an overall asymptomatic positivity rate of 0.67% (weekly positivity rate during this time period ranged from 0.3% to 2.9%). Our automated, high-throughput protocol has enabled us to achieve this scale with a lightly staffed laboratory of ∼6 personnel. Here, we describe the innovations and optimizations that allow us to test tens of thousands of samples a week (**Figure 3**).

**Figure 3.**
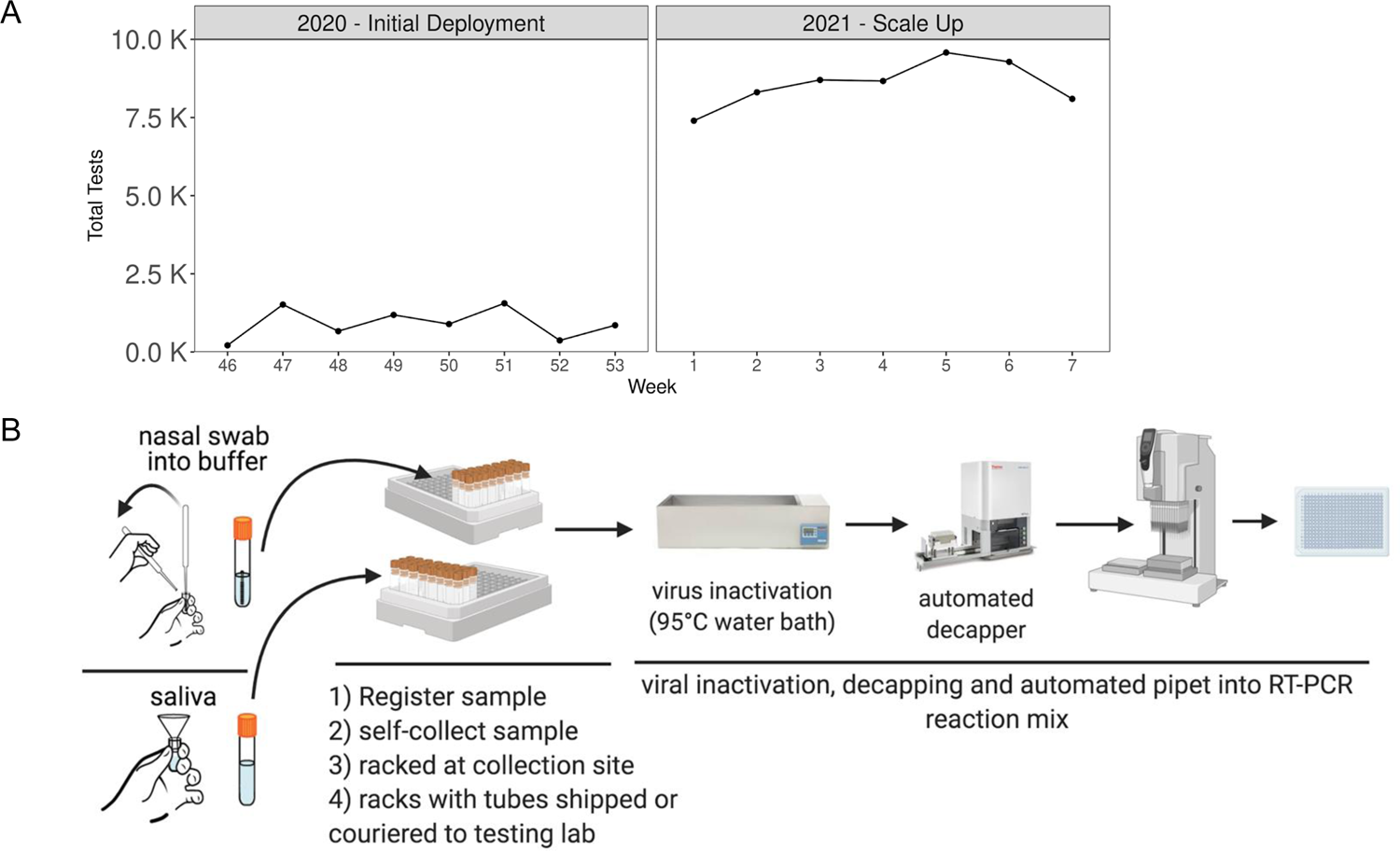
Deployment of Clinical Testing at the UCLA COVID-19 SwabSeq Testing Laboratory. A) Since November 2020, we have used SwabSeq for large-scale screening in conjunction with our saliva and nasal swab collection processes. We have scaled up to nearly 10,000 samples per week and are continuing to increase our capacity. Week number refers to the week of the year spanning 2020-2021. Our weekly percent positivity rate ranges between 0.3%-2.4% over this period. B) Our clinical deployment streamlined the pre-analytical testing process such that we receive tubes that are ready to be processed through our testing protocol.

Sample registration is one of the most-labor intensive aspects of obtaining samples. We partnered with PreciseMDX and used their website for online ordering of tests and return of results (**Figure S12**). We have tested both self-collected nasal swab and saliva samples (**Figure 3B**); collection was coordinated by the testing site using kits provided by our lab. A sample is registered to an order when barcodes on the sample tubes are scanned using the PreciseMDX software at the time of collection. In doing so, we eliminated the need for laborious and error-prone manual pipetting of samples into uniform containers for testing.

We further improved the efficiency of testing at scale by requiring subjects to use one of two sample collection kits that are compatible with downstream automation. In one kit, saliva samples are collected by custom funnels into tubes that are then racked in a 96-tube format at the collection site and can be uncapped using automated decappers. This allows us to pipet batches of 96-samples at a time directly into a RT-PCR mix (**Figure 3B, Source Video 1 Pre-PCR.mov**). Our second collection kit uses a nasal swab we designed that has a break-off point just below the tip. Therefore, the sample can be processed with the swab without interfering with our downstream testing processes (**Figure S13**).

Translation of Swabseq from the bench to real-world testing required process optimization. The development of extraction-free protocols prioritized “low-touch” handling. Our inactivation protocol requires simple heating of the specimen tube, minimizing exposure and pipetting of the sample. We limit our use of plastic consumables, and use only one liquid-handler tip per sample, thereby minimizing supply chain challenges. Finally, we moved to single-use, index primer plates to eliminate variability in primer pipetting when this is done manually.

How many samples can be processed? We calculate that a single person can process six 384-well plates per hour using an automated liquid handler. In our COVID-19 testing laboratory, we scaled this to four liquid handlers capable of processing up to 24 plates/hour, which is equivalent to a capacity of 9,216 samples per hour. A laboratory running 24 hours a day could handle over 100,000 samples per day. Furthermore, the post-PCR multiplexing and library generation is designed such that a single person can handle thousands of samples concurrently; the number of samples is dependent only on the capacity of the sequencer (**Source Video 2 Post-PCR.mov**). For smaller-scale university settings, testing of up to ∼3000 samples a day would require an automated decapper, a single liquid handler and a MiniSeq, an investment of less than $150,000 in equipment. We provide an R package for fully automated sequence analysis, quality-control reporting, and result interpretation.

These optimizations allow for rapid scaling of the SwabSeq diagnostic platform at the onset of a novel pandemic. We tested sample collection and processing workflows in a variety of settings, including the emergency room, return-to-school testing and university-wide population screening. These and other optimizations demonstrate a path to rapid scaling in population-dense settings such as university campuses.

## DISCUSSION

Swabseq has the potential to alleviate existing bottlenecks in diagnostic clinical testing for Covid19. We believe that it has even greater potential to enable testing on a scale necessary for pandemic suppression via population surveillance. The technology represents a novel use of massively parallel next-generation sequencing for infectious disease surveillance and diagnostics. We have demonstrated that SwabSeq can detect SARS-CoV-2 RNA in clinical specimens from both purified RNA and extraction-free lysates, with clinical and analytical sensitivity and specificity comparable to RT-qPCR performed in a clinical diagnostic laboratory. We have optimized SwabSeq to prioritize scale and low cost, as these are the key factors missing from current COVID-19 diagnostic platforms.

Methods for surveillance testing, such as SwabSeq, should be evaluated differently than those for clinical testing. Clinical testing informs medical decision-making, and thus requires high sensitivity and specificity. For surveillance testing, the important factors are the breadth and frequency of testing and the turn-around-time^12^. Sufficiently broad and frequent testing with rapid return of results, contact tracing, and quarantining of infectious individuals can effectively contain viral outbreaks, avoiding blanket stay-at-home orders. Epidemiological modeling of surveillance testing on university campuses has shown that diagnostic tests with only 70% sensitivity, performed frequently with a short turn-around time, can suppress transmission^13^. However, there remain major challenges for practical implementation of frequent testing, including the cost of testing and the logistics of collecting and processing thousands of samples per day.

The use of next generation sequencing in diagnostic testing has garnered concern about turn-around-time and cost. SwabSeq uses short sequencing runs that read out the molecular indexes and 26 base pairs of the target sequence in as little as 3 hours, followed by computational analysis that can be performed on a desktop computer in 5 minutes. The cost of 1,000 samples analyzed in one MiSeq run is less than $1 per sample for sequencing reagents. Running 10,000 samples on a NextSeq550, which generates 13 times more reads per flow cell, can reduce this sequencing cost approximately 10-fold. We estimate that the total consumable cost, inclusive of the collection kit and informatics infrastructure, ranges from 4 to 6 dollars per test. The total cost of operations depends on a wide range of factors, including the costs of setting up CLIA-certified laboratory, certified personnel, logistics of sample collection, and result reporting. Ongoing optimization to decrease reaction volumes and use less expensive RT-PCR reagents can further decrease the total cost per test.

Finally, scaling up testing for SARS-CoV-2 requires high-throughput sample collection and processing workflows. Manual processes, common in most academic clinical laboratories, are not easily compatible with simple automation. The current protocols with nasopharyngeal swabs into viral transport media, Amies buffer or normal saline are collection methods that date back to the pre-molecular-genetics era, when live viral culture was used to identify cytopathic effects on cell lines. A fresh perspective on collection methods that are easily scalable is enormously beneficial to scaling up centralized laboratory testing approaches.

We have successfully demonstrated that it is possible to set up SwabSeq in a small testing laboratory to process tens of thousands of samples a week, and with appropriate instrumentation to scale to hundreds of thousands. To promote scalability, we have developed sample collection protocols that use smaller-volume tubes compatible with simple automation, such as automated capper-decappers and 96-head liquid handlers^23, 24^. These approaches decrease the amount of hands-on work required in the laboratory to process tests and enable higher reproducibility, faster turn-around time and decreasing exposure risk to laboratory workers. However most of the challenges to scaling lie in developing interfaces for efficient sample registration, patient information collection and return of results. We have shown one way to overcome these challenges but each laboratory will face unique challenges due to the need to meet whatever regulatory requirements are in place, and the need to integrate with existing systems for handling patient information.

The SwabSeq diagnostic platform complements traditional clinical diagnostics tests^25^, as well as the growing arsenal of point-of-care rapid diagnostic platforms^26^ emerging for COVID-19, by increasing test capacity to meet the needs of both diagnostic and widespread surveillance testing. Looking forward, SwabSeq is easily extensible to accommodate additional pathogens and viral targets. This would be particularly useful during the winter cold and flu season, when multiple respiratory pathogens circulate in the population and cannot be easily differentiated based on symptoms alone. Furthermore, in order to track the increasing number of SARS-CoV-2 variant strains, we can sequence additional viral amplicons to genotype key mutations and further improve test sensitivity (**Figure S22**). We believe that SwabSeq could be rapidly spun up within several weeks to accommodate future novel pathogens with pandemic potential. Surveillance testing is likely to become a part of the new normal as we aim to safely reopen the educational, business and recreational sectors of our society.

### SOFTWARE AND DATA

https://github.com/joshsbloom/swabseq (code for data and figure presented)

https://github.com/joshsbloom/swabseqr (R package for fully automated analysis of patient samples)

https://github.com/octantbio/SwabSeq (code for primer design and cross-reactivity)

The core technology has been made available under the Open Covid Pledge, and software and data under the MIT license (UCLA) and Apache 2.0 license (Octant Inc.).

Optimized Protocols

Table S1 - Equipment List

Table S2 - Precision and Repeatability

Table S3 - Primers

Sample Experimental Design Setup

Sample Barcode Translator

## Data Availability

Software and data are available at the shared github links.
The core technology has been made available under the Open Covid Pledge, and software and data under the MIT license (UCLA) and Apache 2.0 license (Octant).

https://github.com/joshsbloom/swabseq

https://github.com/joshsbloom/swabseqr

https://github.com/octantbio/SwabSeq

## Acknowledgments

We thank Jane Semel. Without her support this work would not have been possible. We also thank the Held Foundation and the Carol Moss Foundation for their support of this project. We thank the UCLA David Geffen School of Medicine’s Dean’s Office for their support, the Fast Grants, Inc for funding of this work. We also thank Lea Starita, Beth Martin, Jase Gehring, Sanjay Srivatsan, Jay Shendure, and the members of the Covid Testing Scaleup Slack for their input, guidance and openness in sharing their processes. This work was supported by funding from the Howard Hughes Medical Institute (to LK) and DP5OD024579 (to VA). IL is supported by T32GM008042. We thank Marlene Berro for her guidance with the FDA EUA201963. We also thank the clinical lab scientists in the UCLA Clinical Microbiology lab for their assistance in collecting and processing the remnant specimens and data and our wonderful staff in the UCLA SwabSeq COVID19 Testing laboratory for deploying our CLIA test. We thank Laura Yost and Alex Martin for their advice and guidance during our scaling process. Biorender.com was used to generate figures with cartoons. We thank all the video games and video game makers that have helped keep our loved ones sane as we spent all our time on SwabSeq. E.M.J, M.G., N.B.L., S.W.S., F.Y., E.M.T., K.M.K., and J.P., and S.K. are employed by and hold equity, J.S.B. consults for and holds equity, and A.R.C holds equity in Octant Inc. which initially developed SwabSeq, and has filed for patents for some of the work here, though they have been made available under the Open Covid License: https://www.notion.so/Octant-COVID-License-816b04b442674433a2a58bff2d8288df.

## Data Availability

Source data for all figures are available on https://github.com/joshsbloom/swabseq. All protocols and primers are available under an Open Covid License https://www.notion.so/Octant-COVID-License-816b04b442674433a2a58bff2d8288df.

## Code Availability

All code can be accessed on https://github.com/joshsbloom/swabseq. An R package to automate the resulting of patient samples can be found at https://github.com/joshsbloom/swabseqr.

## Author Contributions

JSB and VA wrote the manuscript with assistance from CL, JF, LK, EE, EJ, AC, NL, MG, SK. EJ, AC, NL, MG, SS, JSB, SK designed barcodes and performed early testing and analysis of protocols and reagents. CL, YY, YZ, LG, RD, MB provided early guidance and key automation resources. EE, DH, NLP and CK developed the registration webapp and IT infrastructure. LS, CM, MG, EJ, NL, SK, IL, OFB, VA, JSB performed and analyzed experiments. ASB and LP analyzed mis-assignment of index barcodes. VA, OG, SC, EH, GO, BJC collected and processed clinical samples. DM optimized operational protocols and, DM, SK, ML, TL, and EE optimized scale up. EE, LK, JF, CL, YY, YZ, JB provided helpful insight into protocols, software, and development and optimization of our specimen collection and handling. FY, EMT, KMK, JP, and MH developed the diversified S Spike mixture, N1 primers and flu primers.

## METHODS

### Sample Collection

All patient samples used in our study were deidentified. All samples were obtained with UCLA IRB approval. Nasopharyngeal samples were collected by health care providers from individuals whom physicians suspected to have COVID19. Upon deployment of testing, we obtained clinical specimens and tested them under our CLIA license (05D2198285) with a laboratory developed test.

### Creation of Contrived Specimens

For the clinical limit of detection experiments, we pooled confirmed, COVID-19 negative remnant nasopharyngeal swab specimens collected by the UCLA Clinical Microbiology Laboratory. Pooled clinical samples were then spiked with ATCC Inactivated Virus (ATCC 1986-HK) at specified concentrations and extracted as described below. For the clinical purified RNA samples, they were collected as nasopharyngeal swabs and purified using the KingFisherFlex (Thermofisher Scientific) instrument using the MagMax bead extraction. All extractions were performed according to manufacturer’s protocols. For extraction-free samples, we first contrived samples at specified concentrations into pooled, confirmed negative clinical samples and diluted samples in TE buffer or water prior to adding to the RT-PCR master mix.

### Processing of Extraction-Free Saliva Specimens

Direct saliva is collected into a Matrix tube (Thermofisher, 3741-BR) using a small funnel (TWDRer 6565). The saliva samples were collected into a matrix tube and heated to 95℃ for 30 minutes. Samples were then either frozen at −80℃ or processed by dilution with 2X TBE with 1% Tween-20, for final concentration of 1x TBE and 0.5% Tween-20^22^. We also tested 1x Tween with Qiagen Protease and RNA Secure (ThermoFisher), which also works but resulted in more sample-to-sample variability and required additional incubation steps.

### Processing of Extraction-Free Nasal Swab Lysates

All extraction-free lysates were inactivated using a heat inactivation at 56℃ for 30 minutes. Samples were then diluted with water at a ratio of 1:4 and directly added to mastermix. Dilution amounts varied depending on the liquid media that was used. We found that of the CDC recommended media, normal saline performed the most robustly. Viral Transport Media and Amies Buffer showed significant PCR inhibition that was difficult to overcome, even with dilution in water. We recommend placing the swab directly into the diluted TE buffer, which has little PCR inhibition.

### Barcode Primer Design

Barcode primers were chosen from a set of 1,536 unique 10bp i5 barcodes and a set of 1,536 unique 10bp i7 barcodes. These 10 bp barcodes satisfied the criteria that there is a minimum Levenshtein^27^ distance of 3 between any two indices (within the i5 and i7 sets) and that the barcodes contain no homopolymer repeats greater than 2 nucleotides. Additionally, barcodes were chosen to minimize homo- and hetero-dimerization using helper functions in the python API to Primer3^28^.

Specificity of each of our primer pairs was tested against 38 other common respiratory viruses, including MERS and SARS-CoV-1 using BLASTn, and no significant homology (defined as >80% homology) was identified. Additional details and code for primer design and specificity analysis can be found at https://github.com/octantbio/SwabSeq.

### Construction of S and RPP30 *in vitro* standard

RT-PCR was performed using primers shown above on gRNA of SARS-CoV-2 (Twist BioSciences, #1) for construction of a *in vitro* S standard DNA template. RT PCR (FP_1, R) and a second round of PCR (FP_2, R) was performed on HEK293T lysate for construction of an *in vitro* RPP30 standard DNA template. Products were run on a gel to identify specific products at ∼150 bp. DNA was purified using Ampure beads (Axygen) using a 1.8 ratio of beads:sample volume. The mixture was vortexed and incubated for 5 minutes at room temperature. A magnet was used to bind beads for 1 minute, washed twice with 70% EtOH, beads were air-dried for 5 minutes, and then removed from the magnet and eluted in 100 μL of IDTE Buffer. The bead solution was placed back on the magnet and the eluate was removed after 1 minute. DNA was quantified by nanodrop (Denovix).

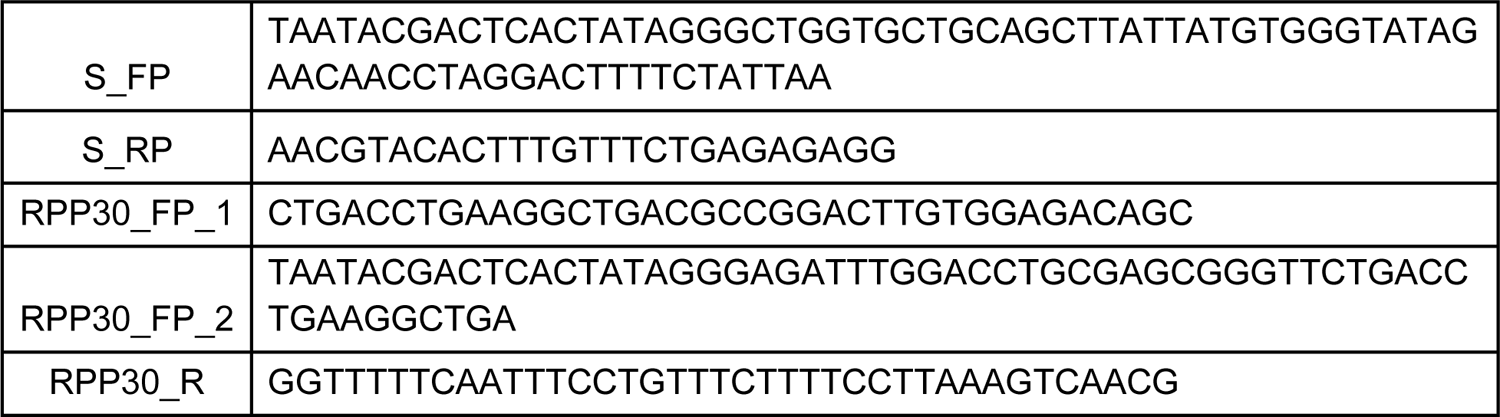

This prepared DNA template was used for standard HiScribe T7 in vitro transcription (NEB). IVT reactions prepared according to the manufacturer’s instructions using 300 ng of template DNA per 20 uL reaction with a 16 hour incubation at 37℃. IVT reactions were treated with DNAseI according to the manufacturer’s instructions. RNA was purified with an RNA Clean & Concentrator-25 kit (Zymo Research) according to the manufacturer’s instructions and eluted into water. RNA standard was quantified both by nanodrop and with a RNA screen tape kit for the TapeStation according to the manufacturer’s instructions (Agilent) to verify the RNA was the correct size (∼133 nt).

### Construction of S-diversified synthetic spike-in standard

The diversified S Spike is a control for reverse transcription and amplification of the viral sequence in each RT-PCR well. The diversified S Spike is designed to have a similar sequence to the S amplicon, but each spike has a unique 26 base pair region that is read by the sequencer that allows for improved base-calling and removes the reliance on PhiX loading. This was achieved by ensuring equal base coverage at each position along the 26 base pair region. This amplicon can be differentiated from the true viral S amplicon by sequencing. The S primer pair will amplify both S amplicon and the S diversified spike, at equivalent efficiency. These RNA oligonucleotides are custom synthesized by Synthego, Inc and supplied as 4 separate tubes of equal concentration that are mixed in equal ratios prior to testing. This equimolar mix of RNA is spiked-in every well prior to RT-PCR.

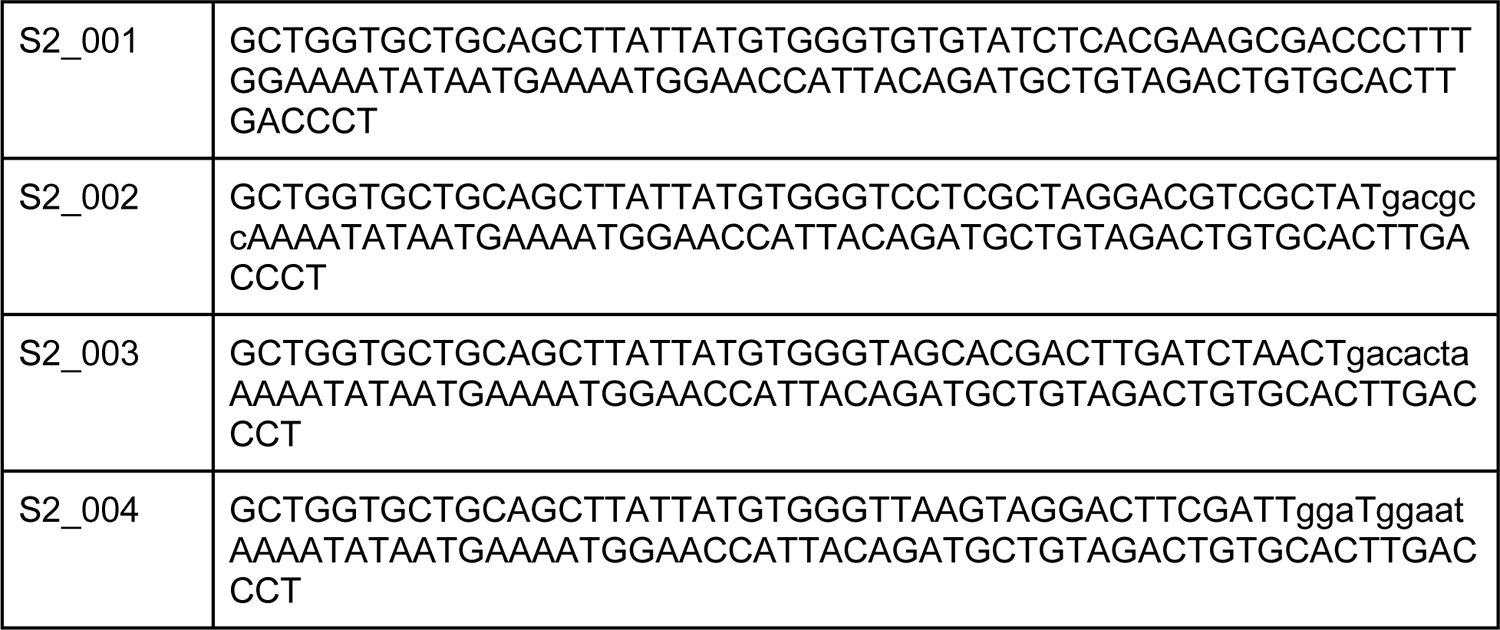

### One-Step RT-PCR

RT-PCR were performed using either the Luna® Universal One-Step RT-qPCR Kit (New England BioSciences E3005) or the TaqPath™ 1-Step RT-qPCR Master Mix (Thermofisher Scientific, A15300) with a reaction volume of 20 μL. Both kits were used according to the manufacturer’s protocol. The final concentration of primers in our mastermix was 50 nM for RPP30 F and R primers and 400 nM for S F and R primers. Synthetic S RNA was added directly to the mastermix at a copy number of 250 to 500 copies per reaction. We recommend 250 copies per reaction for extraction-free samples and 500 copies for extracted samples. Sample was loaded into a 20 μL reaction. All reactions were run on a 96- or 384-well format and thermocycler conditions were run according to the manufacturer’s protocol. We observe significant differences between the amplification of samples from purified RNA versus extraction-free (unpurified swab) samples (**Figure S14**). For purified RNA samples we performed 40 cycles of PCR. For extraction-free samples, we performed endpoint PCR for 50 cycles.

### Multiplex Library Preparation

After the RT-PCR reaction, samples were pooled using a multichannel pipet or Integra Viaflow Benchtop liquid handler. 6 μL from each well were combined in a sterile reservoir and transferred into a 15 mL conical tube and vortexed. 100 uL of the pool was transferred to a 1.7 mL eppendorf tube for a double-sided SPRI cleanup ^29^. Briefly, 50 μL of AmpureXP beads (Beckman Coulter A63880) were added to 100 μL of the pooled PCR volume and vortexed. After 5 minutes, a magnet was used to collect beads for 1 minute and supernatant transferred to a new eppendorf tube. An additional 130 μL of Ampure XP beads were added to the 150 μL of supernatant and vortexed. After an additional 5 minutes, the magnet was used to collect beads for 1 minute and the beads were washed twice with fresh 70% EtOH. DNA was eluted off the beads in 40 μL of Qiagen EB buffer. The magnet was used to collect beads for 1 minute and 33 μL of supernatant was transferred to a new tube. Purified DNA was quantified and library quality was assessed using the Agilent TapeStation. We observe some differences in non-specific peaks in our TapeStation analysis of the final library preparation, particularly when sequencing extraction-free samples out to 50 cycles (**Figure S15**). The presence of non-specific products affects the quantification of the library, loading concentration, and cluster density.

### Sequencing Protocol with original S-standard

Libraries were sequenced on either an Illumina MiSeq (2012), NextSeq550, or MiniSeq. Prior to each MiSeq run, a bleach wash was performed using a sodium hypochlorite solution (Sigma Aldrich, 239305) according to Illumina protocols. We also perform a maintenance wash between each run. On the NextSeq550 and MiniSeq, the post-run wash was performed automatically by the instruments, and no human intervention was required. The pooled and quantitated library was diluted to a concentration of 6 nM (based on Qubit 4 Fluorometer and Illumina’s formula for conversion between ng/μl and nM) and was loaded on the sequencer at either 25 pM (MiSeq), 1.35 pM (NextSeq), or 1.6 pM (MiniSeq). PhiX Control v3 (Illumina, FC-110-3001) was spiked into the library at an estimated 30-40% of the library. PhiX provides additional sequence diversity to Read 1, which assists with template registration and improves run and base quality.

For this application, the MiSeq and MiniSeq (Rapid Run Kit) require 2 custom sequencing primer mixes, the Read1 primer mix and the i7 primer mix. Both mixes have a final concentration of 20 μM of primers (10 μM of each amplicon’s sequencing primer). The NextSeq requires an additional sequencing primer mix, the i5 primer mix, which also has a final concentration of 20 μM. The MiSeq Reagent Kit v3 (150-cycle; MS-102-3001) is loaded with 30 μL of Read1 sequencing primer mix into reservoir 12 and 30 μL of the i7 sequencing primer primer mix into reservoir 13. The Miniseq Rapid High Output Reagent Kit (100-cycle; 20044338) is loaded with 17 μl of Read1 sequencing primer mix into reservoir 24 and 26 μl of i7 sequencing primer mix into reservoir 28. The NextSeq 500/550 Mid Output Kit (150 cycles; 20024904) is loaded with 52 μl of Read1 sequencing primer mix into reservoir 20, 85 μl of i7 sequencing primer mix into reservoir 22, and 85 μl of i5 sequencing primer mix into reservoir 22. Index 1 and 2 are each 10 bp, and Read 1 is 26 bp.

### Sequencing Protocol with optimized diversity S-standard

The pooled and quantitated library was diluted to a concentration of 5 nM (using the High Sensitivity DNA Qubit and Illumina’s formula for conversion between ng/μl and nM assuming 195bp for the length of the library). Without PhiX we observed more consistent sequencing results if we underloaded the sequencers to 83% of the recommended loading concentrations specified by Illumina. Libraries were loaded on the sequencer at either 1.25 pM (NextSeq), or 1.5 pM (MiniSeq). Custom sequencing primer mixes were prepared and loaded onto sequencers as described in the section above, except we load a larger volume of the Read1 primer mix (24.5uL) onto the MiniSeq.

### Diversified N1 spike & primer design

N1 amplicon was designed by using the CDC RT-PCR primer sequences for SARS-CoV-2 (2019-nCoV_N1-F; GACCCCAAAATCAGCGAAAT and 2019-nCoV_N1-R; TCTGGTTACTGCC AGTTGAATCTG). Diversified N1 spikes were designed using the same principles as the S diversified spike design: four spike sequences with conserved homology arm in the primer and Illumina read 1 oligonucleotide binding region but diversified 26 base pair read region ensuring equal base coverage in each sequencing cycle.

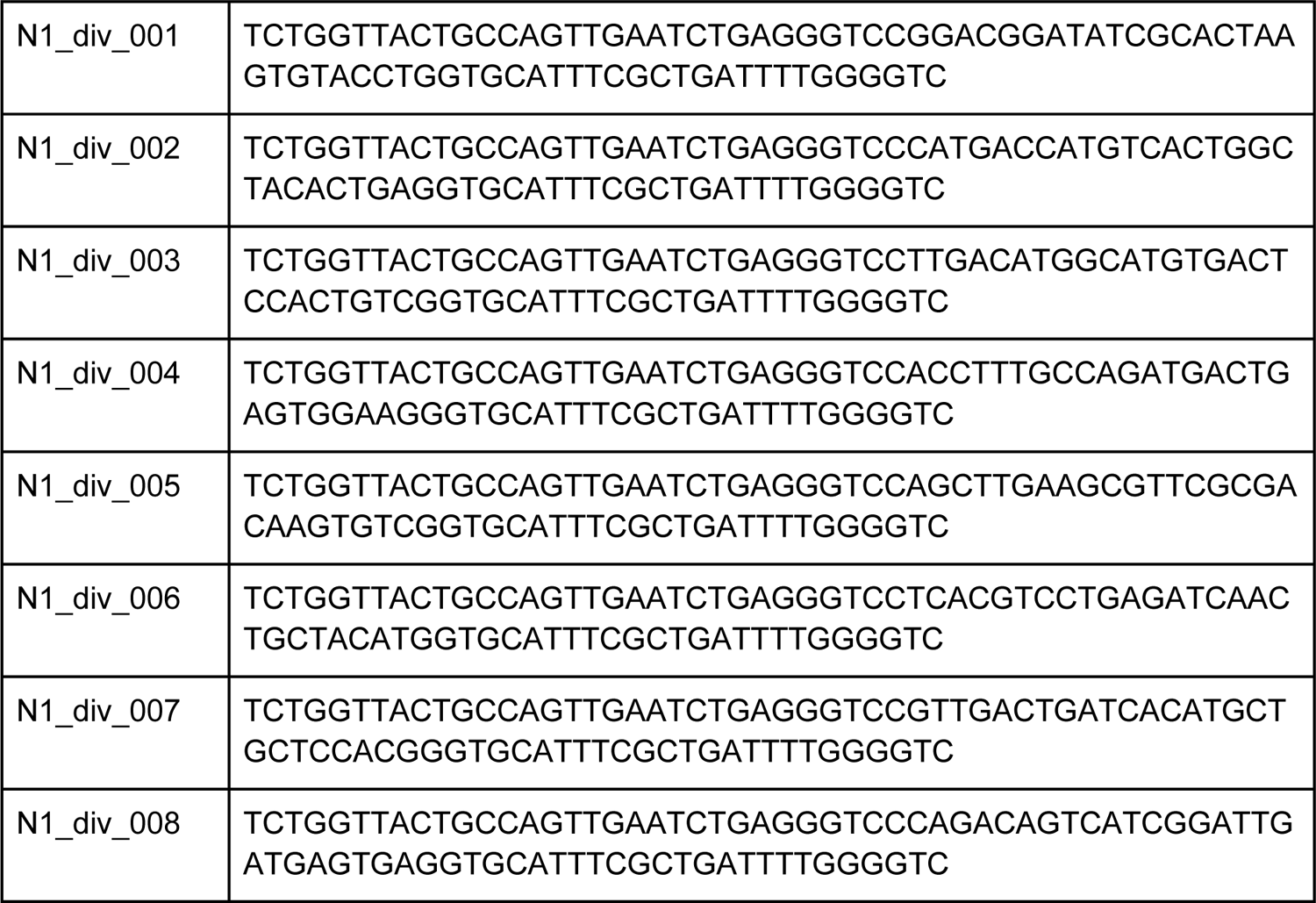

### FluA and FluB primer design

To implement SwabSeq for other respiratory pathogens, we designed universal Influenza A and Influenza B SwabSeq primers. For Influenza A, we designed and tested 2 primer pairs targeting Influenza A M1 gene (FluA_5_f1 GACCAATCYTGTCACCTCTGAC, FluA_5_f2 GACCAATYCTGTCACCTYTGAC, FluA_5_r AGGGCATTTTGGAYAAAGCGTCTA) and NS1 (FluA_12_f TTGGGGTCCTCATCGGAG, FluA_12_r TTCTCCAAGCGAATCTCTGTA) genes.

For Influenza B, we designed and tested a primer pair specific for the NS1 gene (FluB_11_f AAGATGGCCATCGGATCC, FluB_11_r GTCTCCCTCTTCTGGTGATAATC) gene. An in silico cross-reactivity analysis was performed as described for the S and RPP30 primers, none of the pathogens exhibit greater than 80% homology with any of the primers.

### Bioinformatic Analysis

The bioinformatic analysis consists of standard conversion of BCL files into FASTQ sequencing files using Illumina’s bcl2fastq software (v2.20.0.422). Demultiplexing and read counts per sample are performed using our custom software. Here read1 is matched to one of the three expected amplicons allowing for the possibility of a single nucleotide error in the amplicon sequence. The *hamming distance* is the number of positions at which the corresponding sequences are different from each other and is a commonly used measure of distance between sequences. Samples are demultiplexed using the two index reads in order to identify which sample the read originated from. Observed index reads are matched to the expected index sequences allowing for the possibility of a single nucleotide error in one or both of the index sequences. The set of three reads are discarded if both index1 and index2 have hamming distances greater than 1 from the expected index sequences. The count of reads for each amplicon and each sample is calculated. In this analysis we make use of a few custom scripts written in R that rely on the ShortRead^30^ and stringdist^31^ packages for processing fastq files and calculating hamming distances between observed and expected amplicons and indices. This approach was conservative and gave us very low level control of the sequencing analysis. However, we anticipate that continued development of the kallisto and bustools SwabSeq analysis tools^32^ will be a more user-friendly and computationally efficient solution for SwabSeq. We developed an R package (swabseqr) to fully automate sequence analysis, quality-control reporting, and result interpretation.

### Criteria for Classification of RNA-Purified Patient Samples

For our analytic pipeline, we developed QC metrics for each type of specimen. For purified RNA, we require each sample to have at least 10 reads detected for RPP30 and that the sum of S and *in vitro* S standard reads exceeds 2,000 reads. If these conditions are not met, the sample is rerun one time and if there is a second fail we request a resample. To determine if SARS-CoV-2 is present, we calculate if the ratio of S to *in vitro* S standard exceeds 0.003. (We note that we add 1 count to both S and *in vitro* S standard before calculating this ratio to facilitate plotting the results on a logarithmic scale.) If the ratio is greater than 0.003 we concluded that SARS-CoV-2 is detected for that sample and if it is less than or equal to 0.003 we conclude that SARS-CoV-2 is not detected (**Figure S5C**).

The same pair of primers will amplify both the S and *in vitro* S standard amplicons. Because we run an endpoint assay, the primers will be the limiting reagent to continued amplification. In developing this assay, we observed that as S counts increase for a sample, the *in vitro* S standard counts decrease (**Figure S3**). We found that at very high viral levels, *in vitro* S standard read counts decreased to less than 1000 reads. Therefore, analysis of S and *in vitro* S standard together allowed our QC to call SARS-CoV-2 even at extremely high viral levels.

Since the S and *in vitro* S standard are derived from the same primer pair, to account for the scenario where *in vitro* S standard counts are low because S amplicon counts are very high and the sample contains large amounts of SARS-CoV-2 RNA (**Figure S3**) in the QC we require that the sum of S and *in vitro* S standard counts together exceeds 2000. For example, if we detected greater than 2000 S counts and 0 *in vitro* S standard counts this would certainly be a SARS-CoV-2 positive sample and we would result: SARS-CoV-2 detected.

### Criteria for Classification of Extraction Free Patient Samples

We require that the sum of S and S synthetic spike-in reads exceeds 500 reads, or the results are considered inconclusive. With inhibitory lysates we have observed that this lower total is acceptable for maintaining sensitivity and specificity while limiting the number of tests that are considered inconclusive. If the sum of S and S synthetic spike-in reads exceeds 500, we determine if SARS-CoV-2 is detected in a sample by seeing if the ratio of S to S spike exceeds 0.05. (We note that we add 1 count to both S and S spike before calculating this ratio to facilitate plotting the results on a logarithmic scale.) If the ratio is greater than 0.05 we concluded that SARS-CoV-2 is detected for that sample and if it is less than or equal to 0.05 we conclude that SARS-CoV-2 is not detected as long as 10 reads are detected for RPP30 for that sample. This serves as a control that sample collection took place properly and contains a human specimen. If fewer than 10 reads are detected for RPP30 and the ratio of S to S spike is less than or equal to 0.05 the results are considered inconclusive. We have modified this criteria such that only samples without SARS-CoV-2 signal are considered inconclusive if RPP30 reads are fewer than 10. This ensures that we do not miss SARS-CoV-2 positive samples that may have fewer RPP30 reads (**Figure S8**).

### Downsampling analysis

Reads were downsampled from the results for the NP purified confirmatory LoD shown in (**Figure S5B**). We observed that downsampling down to 5,000 reads per well resulted in no instances of mis-classification of SARS-CoV-2 presence or absence. At 5,000 reads per well approximately 3% of wells would no longer pass the filter that the sum of S and S spike reads exceeded 1,000 reads and would result in a sample being classified as ‘Inconclusive’. A logistic regression classifier described elsewhere^19^ should robustly tolerate a small fraction of outlier samples with slightly lower read depth.

### Analysis of index mis-assignment

Unique dual indices and amplicon specific indices were used to study index mis-assignment. In this scheme, each sample was assigned two unique indices for the S or Spike amplicon and two unique indices for the RPP30 amplicon for a total of four unique indices per sample. A count matrix with all possible pairwise combinations, i.e. a “matching matrix”, was generated for each index pair (one i7 and one i5) using kallisto and bustools ^33^. The counts on the diagonal of the matching matrix correspond to input samples and counts off of the diagonal correspond to index swapping events. The extent of index mis-assignment for the i7 and i5 index was determined by computing the row and column sums, respectively, of the off-diagonal elements of the matching matrix. The observed rate of index swapping to wells with a known zero amount of viral RNA was determined by computing the mean of the viral S counts to spike ratio for those wells.

## Supplemental Results

### Improving Limit of Detection Requires Minimizing Sources of Noise

One of the major challenges in running a highly sensitive molecular diagnostic assay is that even a single contaminant or source of noise can decrease the test’s analytical sensitivity. In the process of developing SwabSeq, we observed S reads from control samples in which no SARS-CoV-2 RNA was present (**Figure 1D**). We subsequently refer to these reads as “no template control” (NTC) reads. A key part of SwabSeq optimization has been understanding and minimizing the sources of NTC reads in order to improve the limit of detection (LoD) of the assay. We identified two important sources of NTC reads: molecular contamination and mis-assignment of sequencing reads.

To minimize molecular contamination, we followed protocols and procedures that are commonly used in molecular genetic diagnostic laboratories^34^. To limit molecular contamination, we use a dedicated hood for making dilutions of the synthetic RNA controls and master mix. At the start of each new run, we sterilize the pipettes, dilution solutions, and PCR plates with 10% bleach, followed by UV-light treatment for 15 minutes.

To prevent post-PCR products that are at high concentration from contaminating our pre-PCR processes, we physically separated pre- and post-PCR steps of our protocol into two separate rooms, where any post-PCR plates were never opened within the pre-PCR laboratory space. To further protect from post-PCR contamination, we compared RT-PCR mastermixes with or without Uracil-N-glycosylase (UNG). The presence of UNG in the TaqPath™ 1-Step RT-qPCR Master Mix (Thermofisher Scientific) showed a significant improvement reducing post-PCR contamination of S reads present in the negative patient samples as compared with the Luna One Step RT-PCR Mix (New England Biosciences) (**Figure S16**). The RT-PCR mastermix contains a mix of dTTP and dUTP such that post-PCR amplicons are uracil containing DNA. These post-PCR that are remnants of previously run SwabSeq experiments therefore can be selectively eliminated by UNG. Importantly, this addition does not interfere with downstream sequencing.

A third source of molecular contamination was carryover contamination on the sequencer template line of the Illumina MiSeq^35^. Without a bleach maintenance wash, we found that indices from the previous sequencing run were identified in a subsequent experiment where those indices were not included. While the number of reads for some indices were present at a number of S reads, the presence of carryover contamination affects the sensitivity and specificity of our assay. After an extra maintenance and bleach wash, we substantially reduced the amount of carryover reads present to less than 10 reads (**Figure S17**).

Another source of NTC reads is mis-assignment of amplicons. Mis-assignment of amplicons occurs when sequencing (and perhaps at a lower rate, oligo synthesis) errors result in an amplicon sequence that originates from the *in vitro* S standard but is mistakenly assigned to the S sequence within a given sample. Only 6 bp distinguishes S from *in vitro* S standard at the beginning of read 1. Sequencing errors can result in *in vitro* S standard reads being misclassified as S reads as error rates appear to be higher in the beginning of the read (**Figure S18A**). If computational error correction of the amplicon reads is too tolerant, these reads may be inadvertently counted to the wrong category. To reduce this source of S read misassignment, we use a more conservative thresholding on edit distance (**Figure S18B**). Future redesigns or extensions to additional viral amplicons should consider engineering longer regions of sequence diversity here.

An additional source of NTC reads is when S amplicon reads are mis-assigned to the wrong sample based on the indexing strategy. In our assay, individual samples are identified by pairs of index reads (**Figure 1B**). Mis-assignment of samples to the wrong index could occur if there is contamination of index primer sequences, synthesis errors in the index sequence, sequencing errors in the index sequences, or “index hopping” ^36^.

We leveraged multiple indexing strategies in our development of SwabSeq, from fully combinatorial indexing (where each possible combination of i5 and i7 indices was used to tag samples in the assay) to unique-dual indexing (UDI) where each sample has distinct and unrelated i7 and i5 indices (**Figure S19**). However, the ability to scale can be limited due to the substantial upfront cost of developing that many unique primers. Fully combinatorial indexing approaches significantly expand the number of unique primer combinations. We have also explored a compromise strategy between fully combinatorial indexing and UDI where sets of indices are only shared between small subsets of samples. Such designs reduce the effect of sample mis-assignment while facilitating scaling to tens of thousands of patient samples (**Figure S19**). With a fully combinatorial indexing (**Figure S19**) we observed that NTC read depth was correlated with the total number of S reads summed across all samples that shared the same i7 sequence (**Figure S20A**). This is consistent with the effect of index hopping from samples with high S viral reads to samples that share the same indices. It is possible to computationally correct for this effect, for example using a linear mixed model (**Figures S20B**).

Finally, the challenges associated with combinatorial and semi-combinatorial indexing strategies can be mitigated by using unique dual indexing (UDI), a known strategy to reduce the number of index-hopped reads by two orders of magnitude^37^. We have observed consistently lower S viral reads for negative control samples with this strategy. It also enables quantification of index mis-assignment by counting reads for index combinations that should not occur in our assay (**Figure S21A and B**). The number of index hopping events is correlated with the total number of S + S spike reads (**Figure S21C and D**), indicating that hopped reads are more likely to come from wells where the expected index has strong viral signal. We quantify the overall rate of hopping as 1-2% on a MiSeq, and expect this rate may be higher on patterned flow cell instruments.

There are many sources of noise in amplicon-based sequencing, from environmental contamination in the RT-PCR and sequencing steps to misassignment of reads based on computational correction and “index-hopping” on the Illumina flow cells. Preventing and correcting these sources of error considerably improves the limit of detection of the SwabSeq assay.

### Extension of SwabSeq to additional amplicons in SARS-Co-V2 and to Influenza A/B

We have designed and tested amplicons as extensions of SwabSeq. The motivation behind these additions are that SARS-CoV-2 continues to evolve and could potentially develop mutations that impact the binding sites of our primer pairs and ultimately decrease the sensitivity of our assay. In addition, the utility of our assay beyond the SARS-CoV-2 pandemic requires additional other respiratory viruses that may be concurrently circulating in the population. We have provided the designs for N1 amplicon and for an N1-diversified Standard design (see Methods, **Table S3**). Our initial validations show that the N1 primer has a limit of detection of 2000 copies/mL in saliva (**Figure S22A**) demonstrating similar detection sensitivity as our S primer. Additional validations for multiplexing primers are ongoing.

Influenza A/B (**see Methods, Table S3**) represent further extension of our assay beyond SARS-CoV-2. As the clinical presentation of COVID-19 is similar to other respiratory viral pneumonia, distinguishing the different viral etiologies will allow for early, precise and preventative treatment of infected patients. We show that these primers are detectable by sequencing assays (**Figure S22B**).

## Supplementary Documents

Optimized Protocol.docx Table S1_Equipment List.xlsx Table S2_PrecisionandReproducibility.xlsx Table S3_Primers.xlsx Sample Experimental Design Setup.xlsx Sample Barcode Translator.xlsx

### Supplementary Figures

Supplementary Figures.pdf

### Supplementary Videos

1_PrePCR.mov

2_PostPCR.mov

---

**Figure S1.**
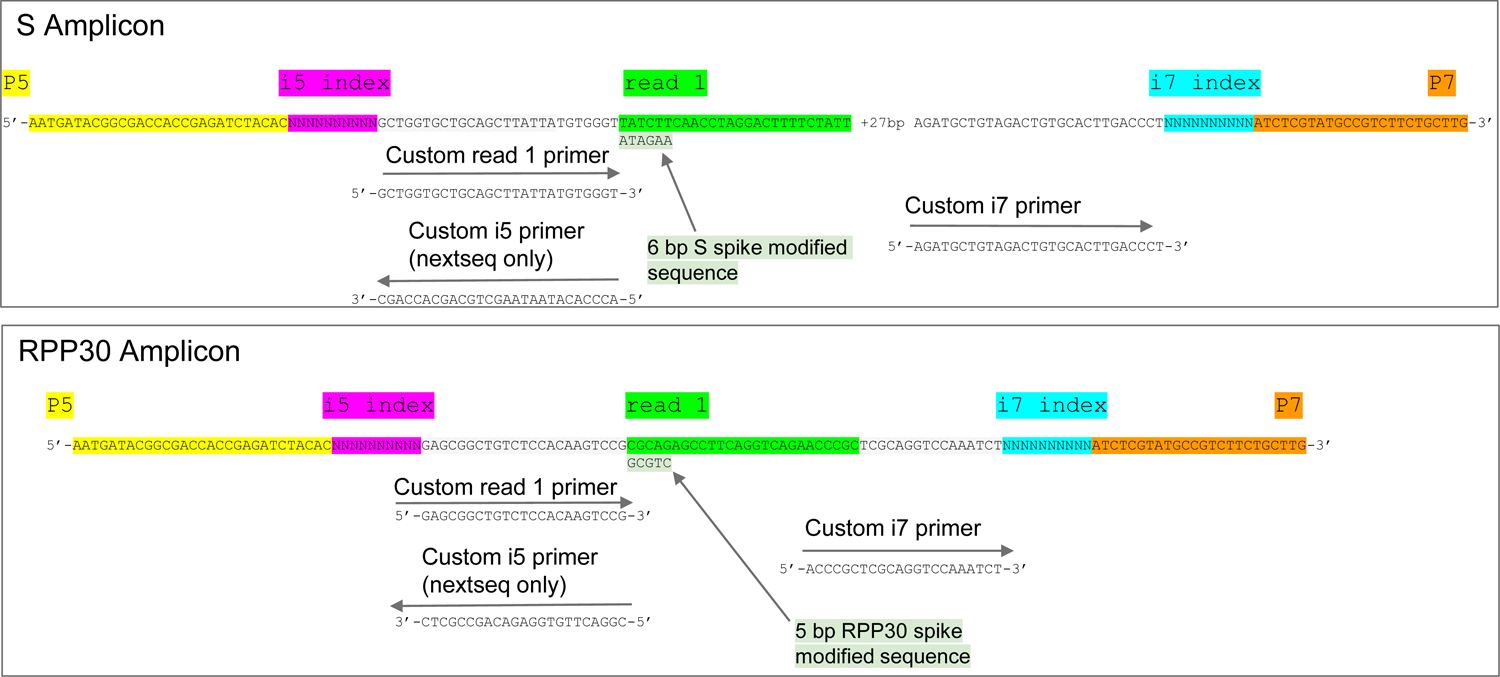
Sequencing library design. The amplicon designs are shown for the S (top) and RPP30 (bottom) amplicons. Amplicons were designed such that the i5 and i7 molecular indexes uniquely identify each sample. SwabSeq was designed to be compatible with all Illumina platforms.

**Figure S2.**
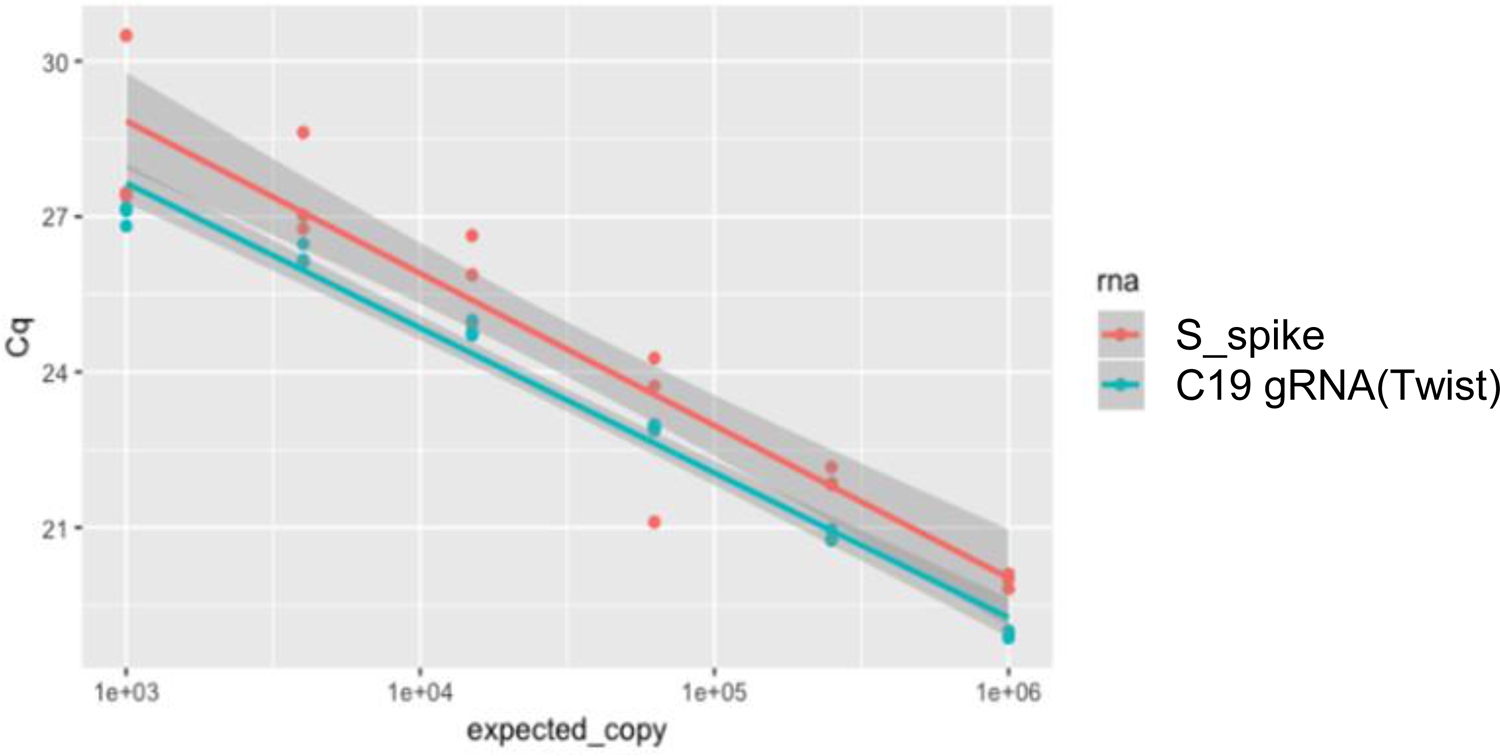
The S primers show equivalent PCR efficiency when amplifying the COVID-19 S gene amplicon and the synthetic *in vitro* S standard. Slope of PCR efficiency of the primers with either the S_spike (labeled in red) or the SARS-CoV-2 viral (labeled in green as C19gRNA) input are as follows: S_spike slope = −6.68e-6 and C19gRNA (Twist Control) slope = −6.74e-6. The slopes are expected to equivalent (parallel) if the primers do not show preferential amplification of the S spike RNA versus the C19gRNA. This shows that the S spike and C19gRNA have equivalent amplification efficiencies using the S primer pair. The bands represent 95% confidence intervals for predicted values, are non-overlapping due to different intercepts, and are not relevant for this analysis of slopes.

**Figure S3.**
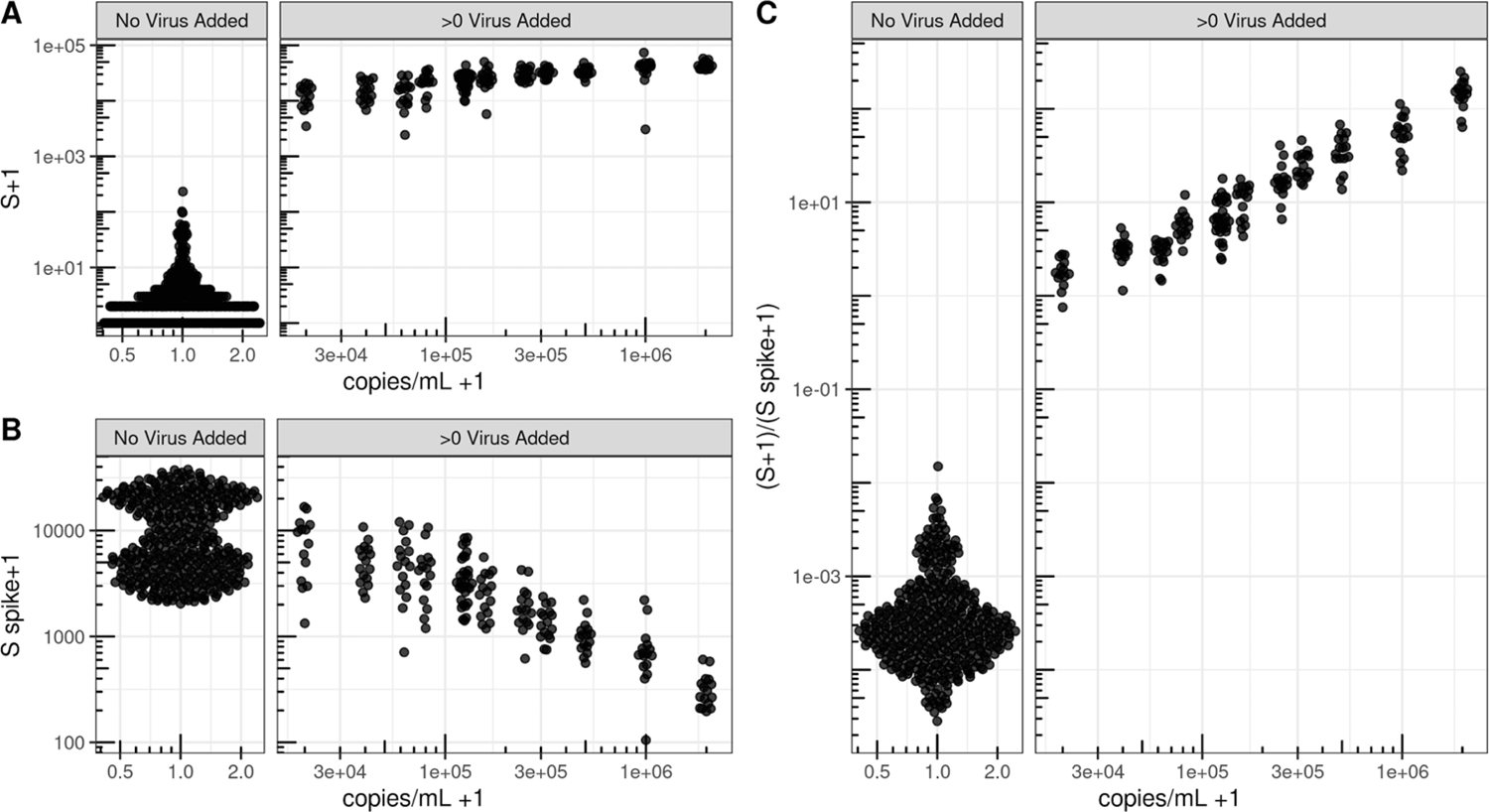
At very high viral concentrations SwabSeq maintains linearity. We include an internal well control, the S Spike, to enable us to call negative samples, even in the presence of heterogeneous sample types and PCR inhibition. (A) As virus concentration increases, we observe increased reads attributed to S and (B) decreased reads attributed to the S Spike. (C) The ratio between the S and S Spike provides an additional level of ratiometric normalization and exhibits linearity up to at least 2 million copies/mL of lysate. Note that ticks on both axes are spaced on a log10 scale.

**Figure S4.**
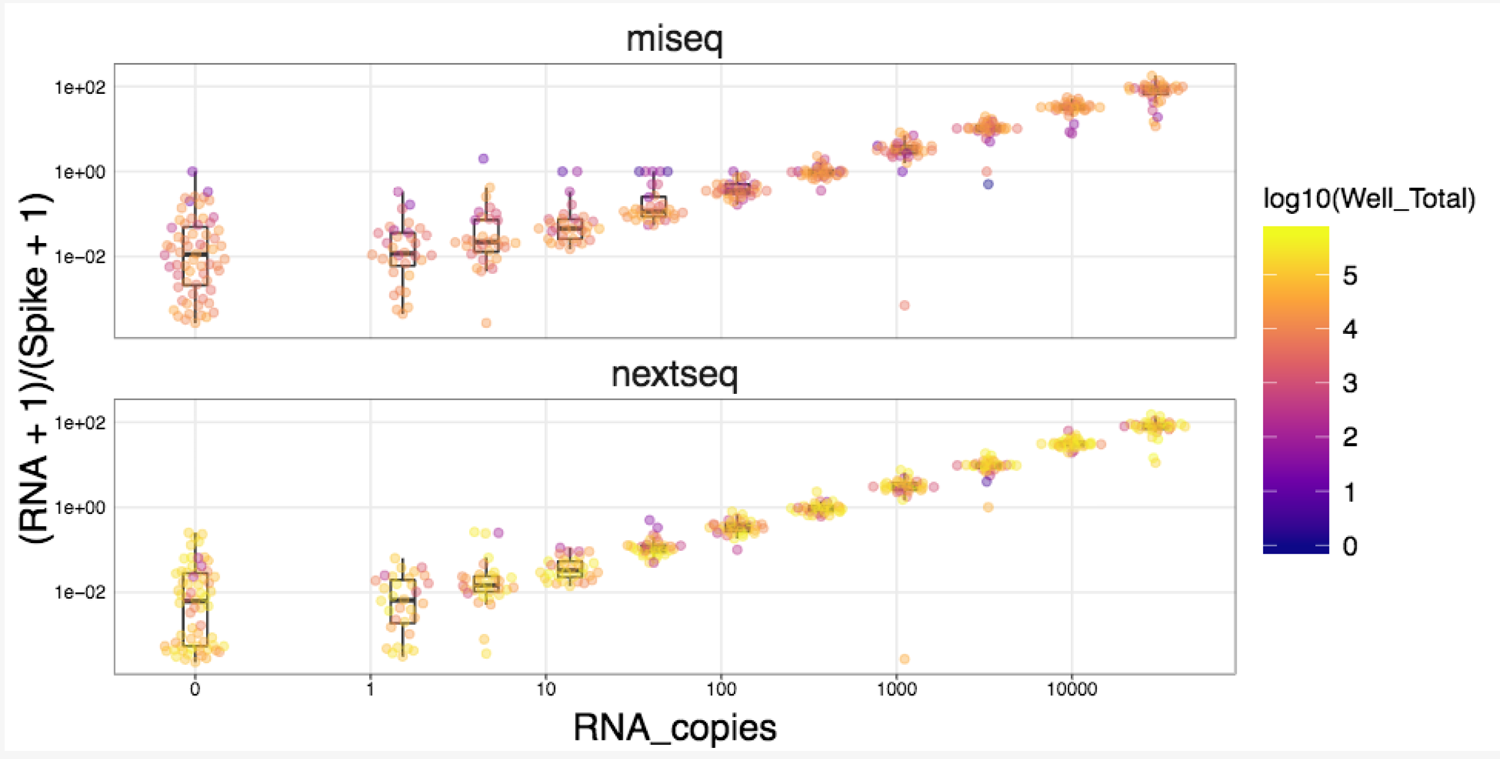
Sequencing is performed on MiSeq or NextSeq Machine with similar sensitivity. Multiplexed libraries run on both MiSeq and NextSeq showed linearity across a wide range of SARS-CoV2 virus copies in a purified RNA background.

**Figure S5.**
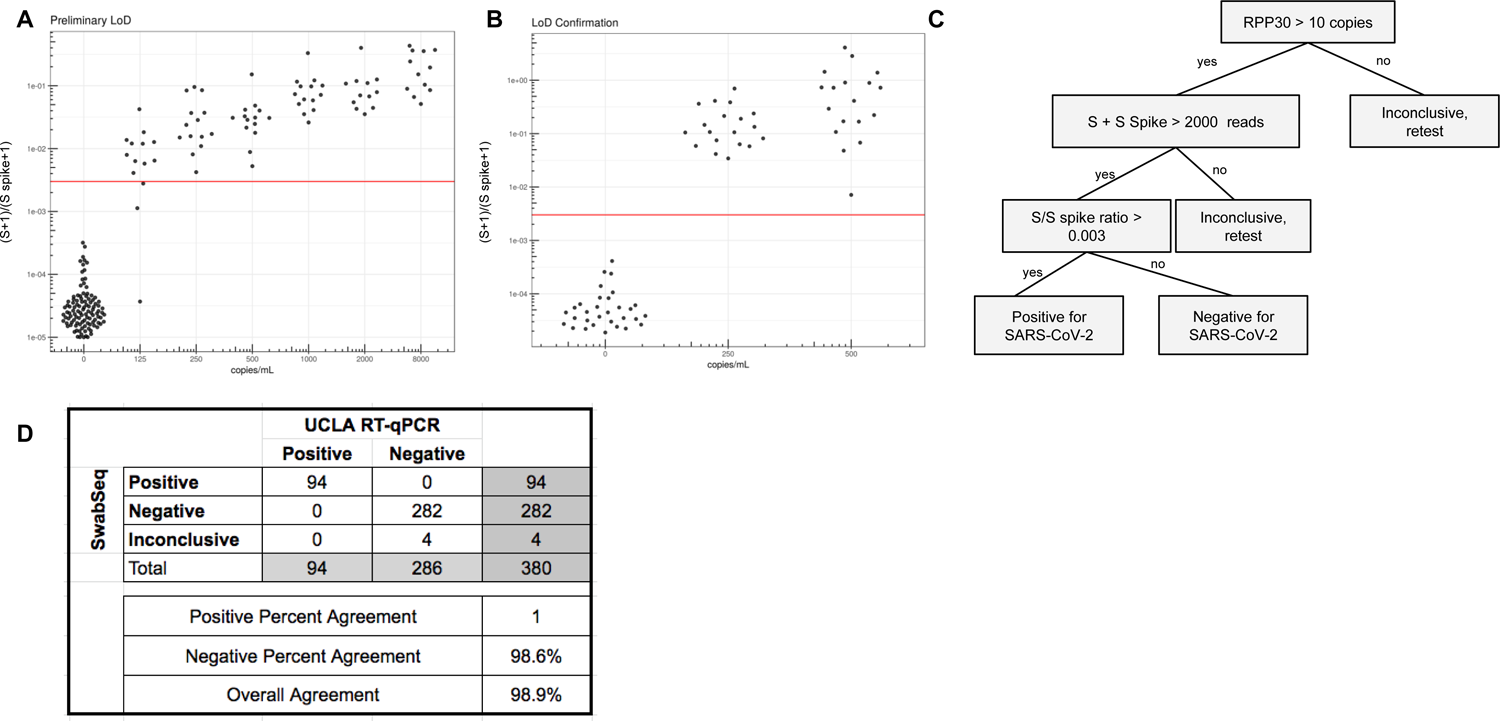
Preliminary and Confirmatory Limit of Detection Data for RNA purified Samples. A) Our preliminary LOD data identified a LOD of 250 copies/mL using the NextSeq550 B) Confirmatory studies showed an LOD of 250 copies/mL using the NextSeq550. C) Our result interpretation guidelines for purified RNA. D) Concordance of the 380 clinical samples that were run during our validation process. Concordance is 98.6%, with 100% positive percent agreement (PPA) and 100% negative percent agreement (NPA).

**Figure S6.**
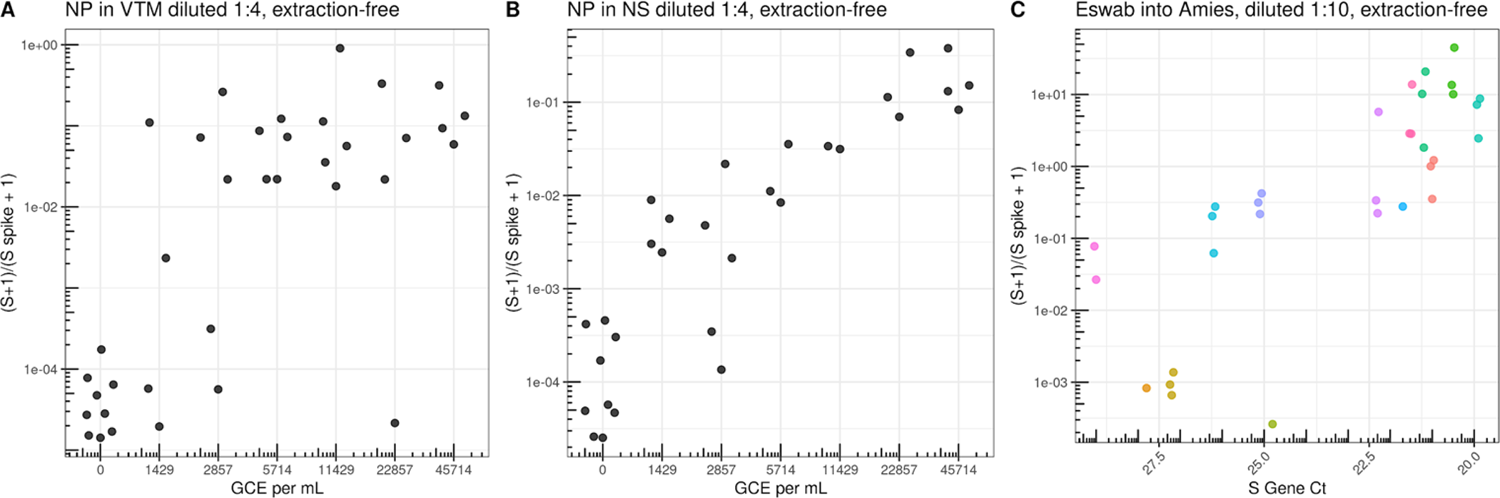
Extraction-Free protocols using traditional collection medias and buffers require dilution to overcome effects of RT and PCR inhibition. A) We tested extraction free protocols for nasopharyngeal swabs that were placed into viral transport media (VTM). We spiked ATCC live inactivated virus at varying concentrations into pooled VTM and then diluted samples 1:4 with water before adding to the RT-PCR reaction. We observed a limit of detection of 5714 copies per mL. B) We also tested nasopharyngeal (NP) swabs that were collected in normal saline (NS), pooled and then spiked with ATCC live inactivated virus at varying concentrations. Contrived samples were diluted 1:4 in water. Here, our early studies show a similar similar limit of detection between 2857 and 5714 copies per mL. C) We tested natural clinical samples that were collected into Amies Buffer (ESwab). Here we compare S gene Ct count (x-axis) from positive samples to the SwabSeq S to S spike ratio (y-axis). Samples were run in triplicate (colors). We observed high concordance for Ct counts of 27 and lower but more variability for Ct counts greater than 27 suggesting that RT and PCR inhibition were affecting our limit of detection. Based on these data we opted only to further explore extraction free protocols into normal saline or tris-EDTA buffer.

**Figure S7.**
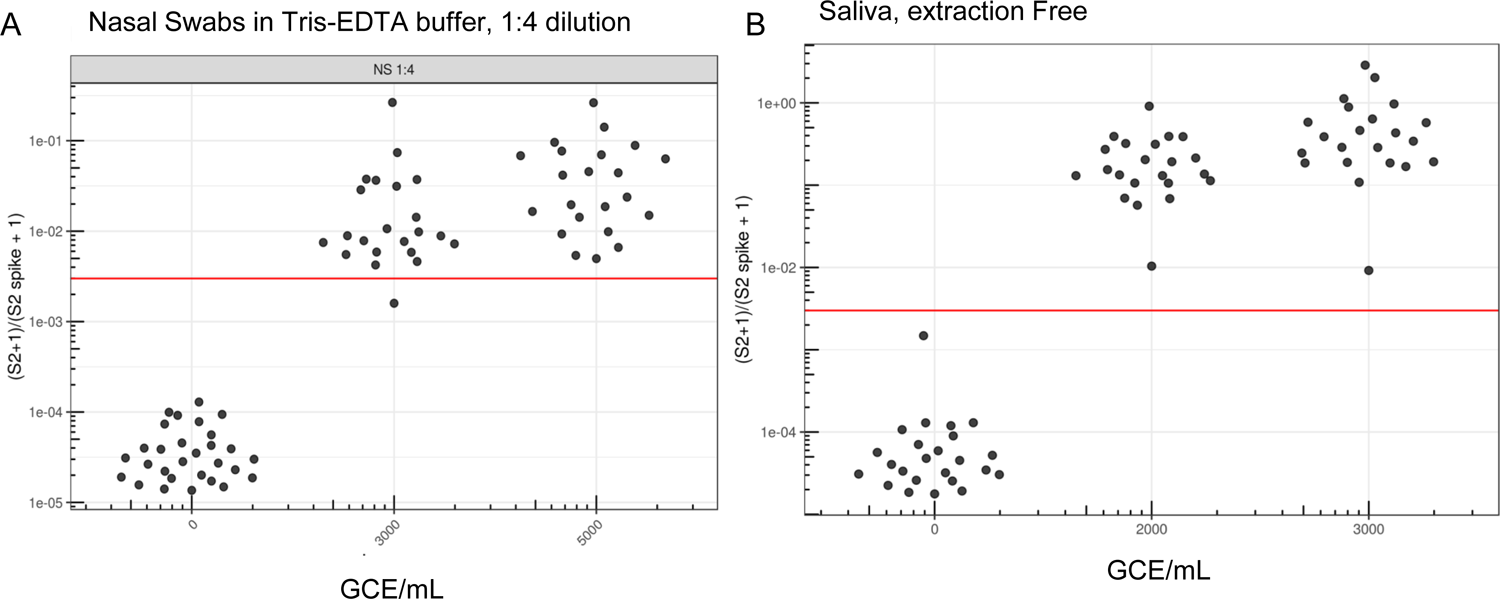
Confirmatory Limit of Detection Data for Extraction Free specimens. A) Our data showed a LOD of 3000 copies/mL for Nasal Swabs samples that were diluted 1:4 in water. These dilutions were performed in replicate at 20 samples per concentration. We tested multiple replicates around the limit of detection. B) Confirmatory studies for extraction-free saliva samples showed an LOD of 2000 copies/mL. Red line indicates the threshold for positivity.

**Figure S8.**
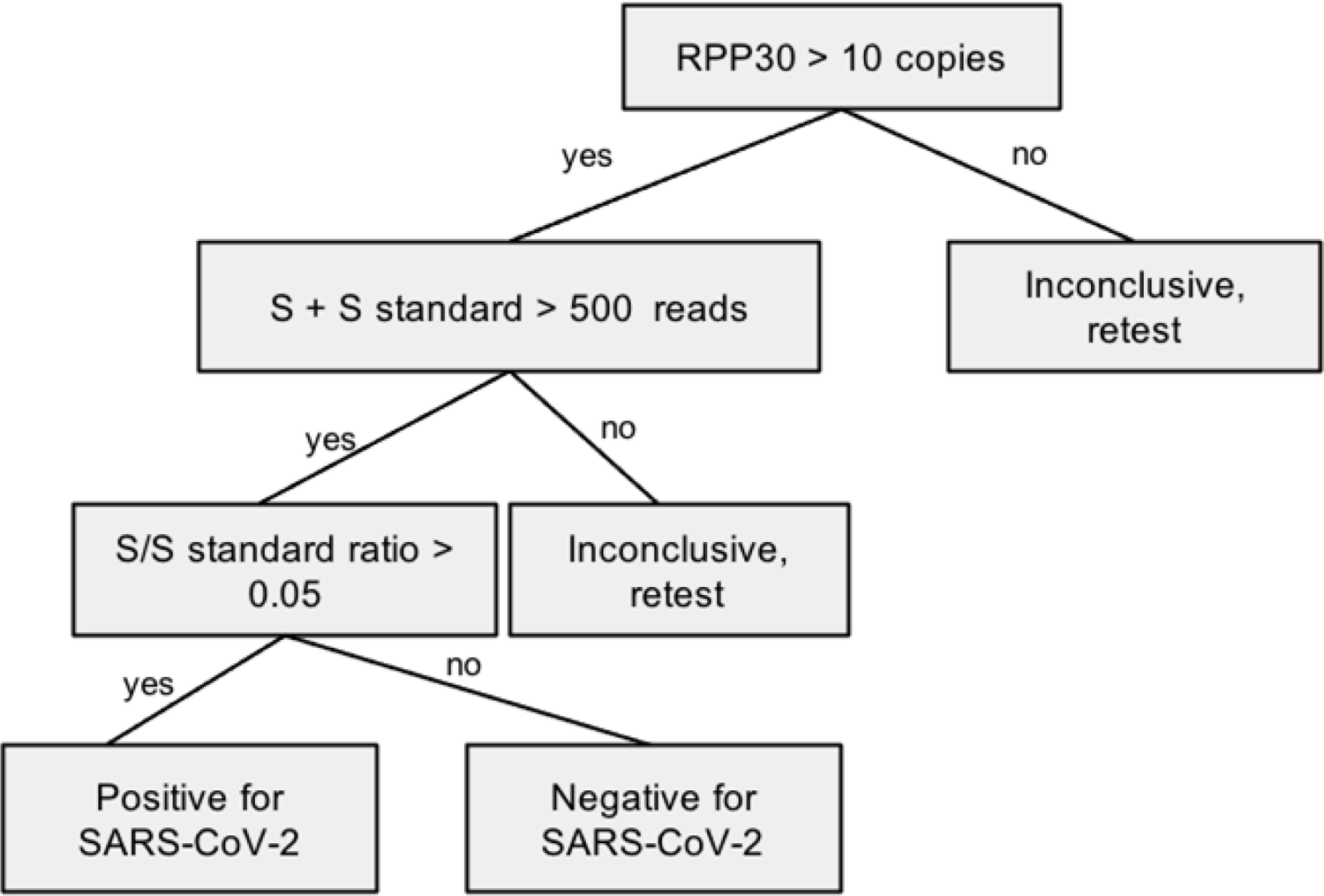
Per sample decision tree for extraction-free samples. Given the slight modifications for our extraction free protocols, we have modified our decision tree to reflect the differences in extraction-free sample types. Our result interpretation guidelines for extraction-free samples relax threshold for S + S spike to 500 reads due to the increased PCR inhibition observed in extraction free sample types. The standard used in our early validations had a slightly lower S/S standard ratio of 0.03 which ultimately in clinical testing had too many false positives. Our current validated test uses the above ratio for extraction free samples.

**Figure S9.**
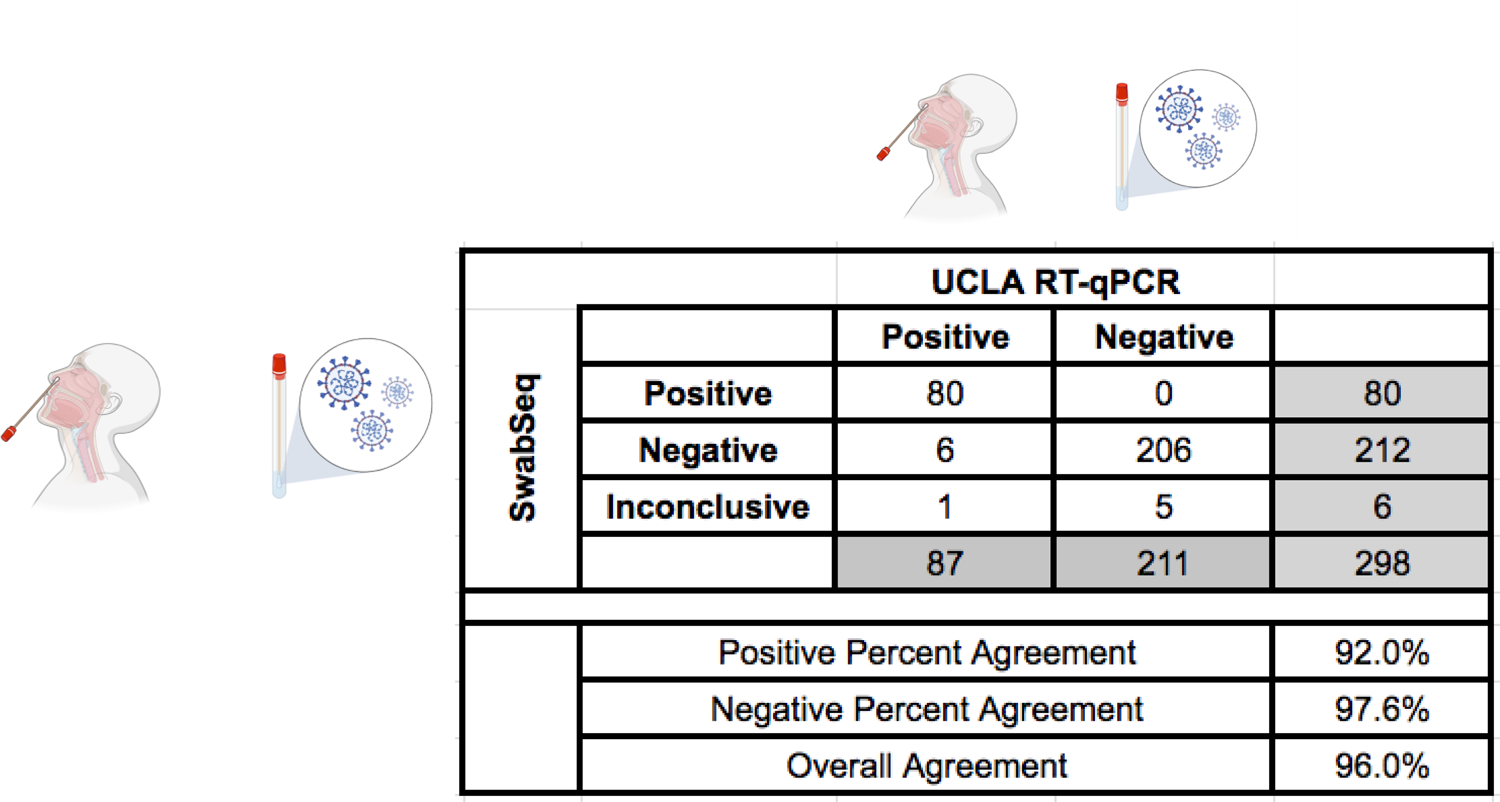
Comparison of extraction-free NP samples run on SwabSeq to NP Swab samples processed to Clinical pathway using RNA purification and RT-qPCR. Evaluation of extraction free nasal swabs processed into normal saline or Tris-EDTA ph 8.0 that have previously tested positive or negative in the UCLA Clinical Microbiology Laboratory. We have explored the sources of false negatives in our data set. Three of the four false negatives stem from differences in the limit of detection, where we do not always detect samples with Ct > 30.

**Figure S10.**
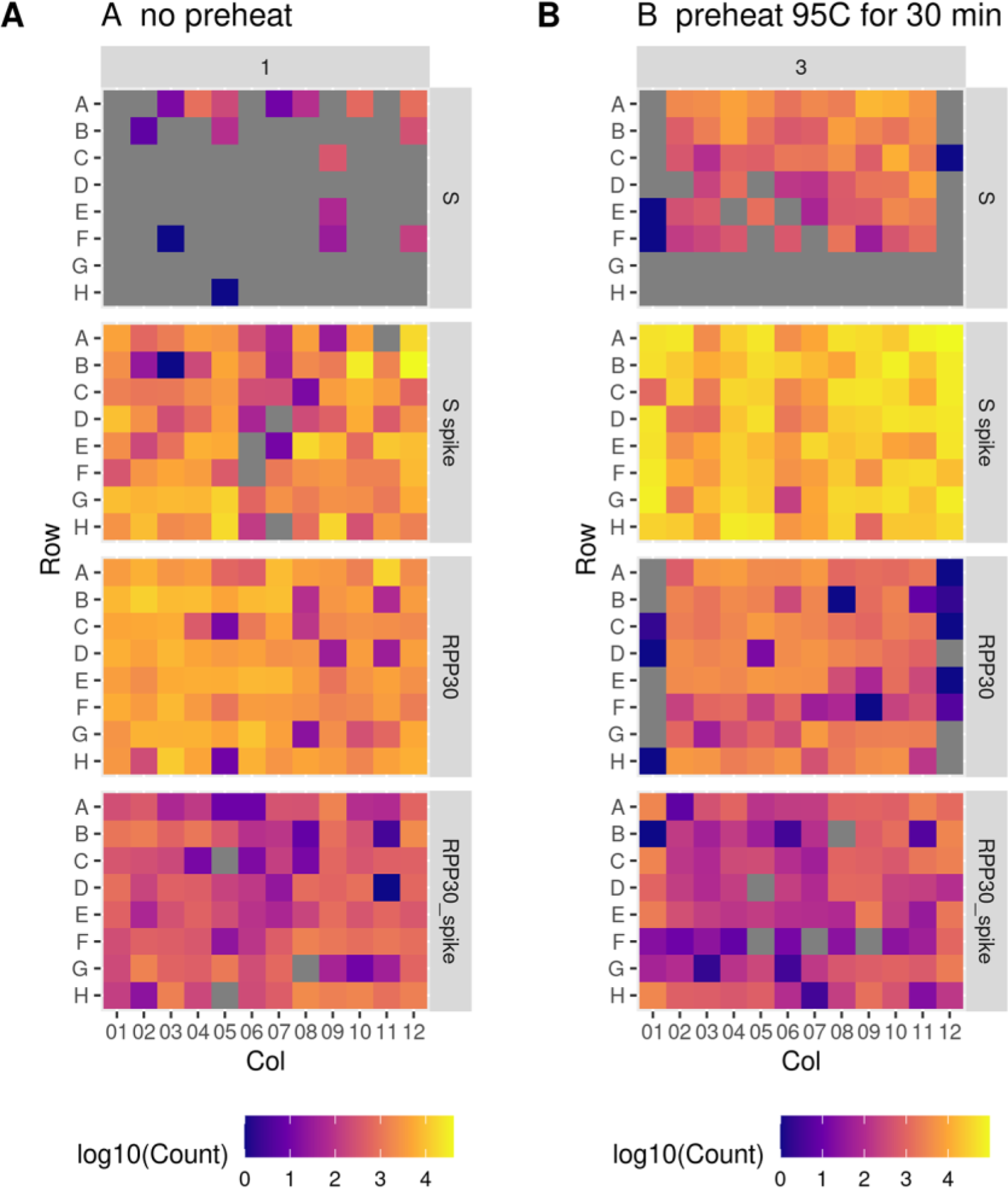
Preheating Saliva to 95C for 30 minutes drastically improves RT-PCR. Detection of viral genome and shows improved robustness in detection of our controls. A) Without preheating, detection of S spike is minimal and there are lower counts for the control amplicons. B) with a 95C preheating step for 30 minutes, we observe robust detection of the S amplicon and synthetic S Spike.

**Figure S11.**
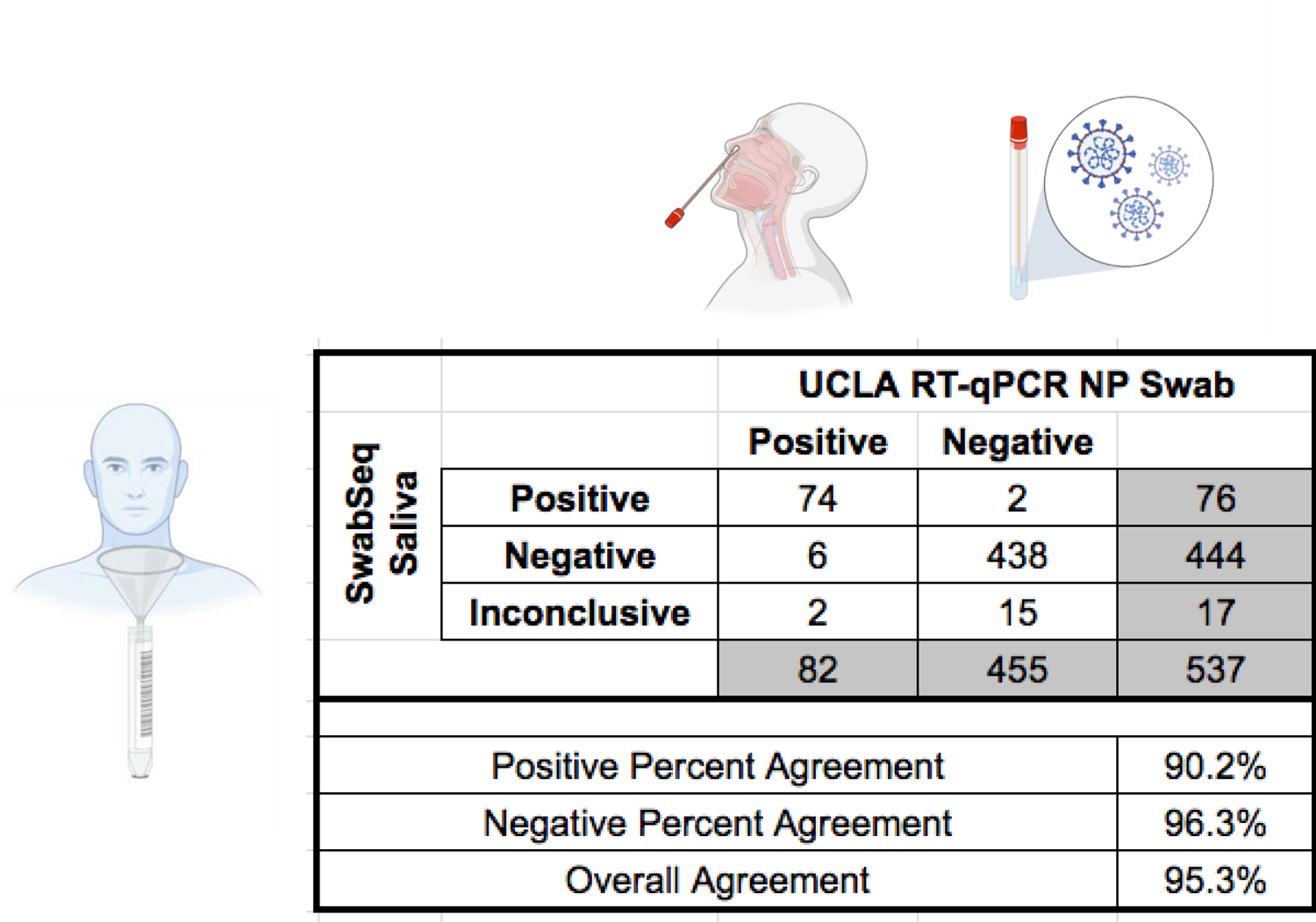
Comparison of extraction-free saliva samples run on SwabSeq to NP Swab samples processed to Clinical pathway using RNA purification and RT-qPCR. We performed a series of studies to compare the concordance of Saliva and NP swab performed within 2 hours of each other. These collections were obtained in the UCLA ED and UCLA Student Health Center over the course of several months.

**Figure S12.**
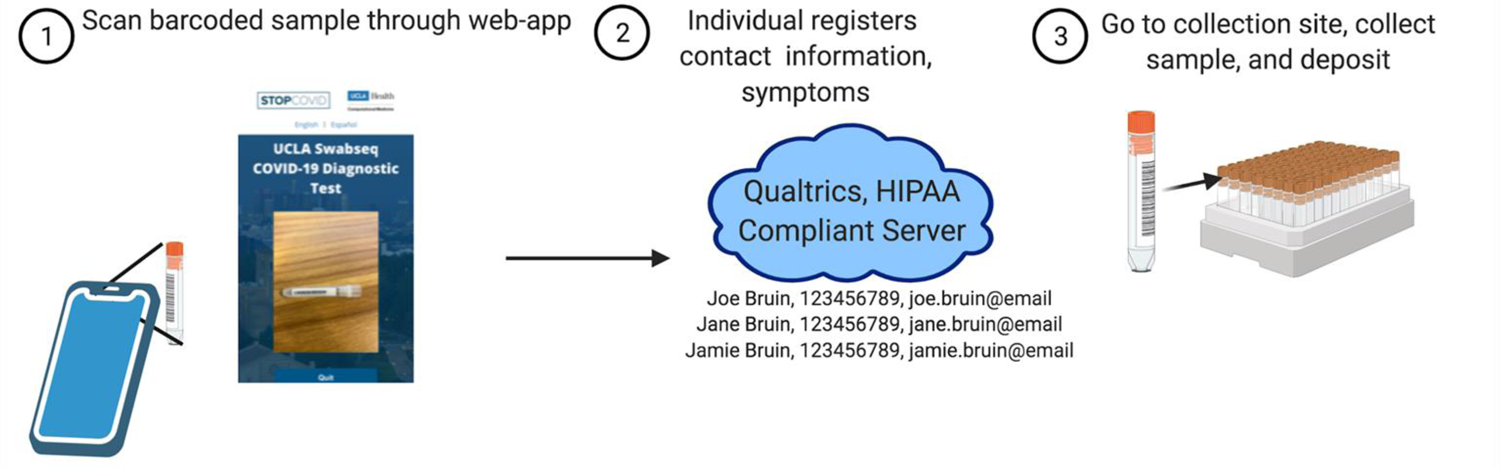
Developing a lightweight sample accessioning to allow for scalable testing into the thousands of samples per day. A major bottleneck is the sample registration to an individual patient. To facilitate the sample accessioning we developed a web-based app for individuals to register their sample tube using a barcode reader and send their identifying information into a secure instance of Qualtrics. In scaled clinical testing in our CLIA laboratory, we used an instance of PreciseQ MDX that allowed organizations to invite cohorts for testing based on their specific needs.

**Figure S13.**
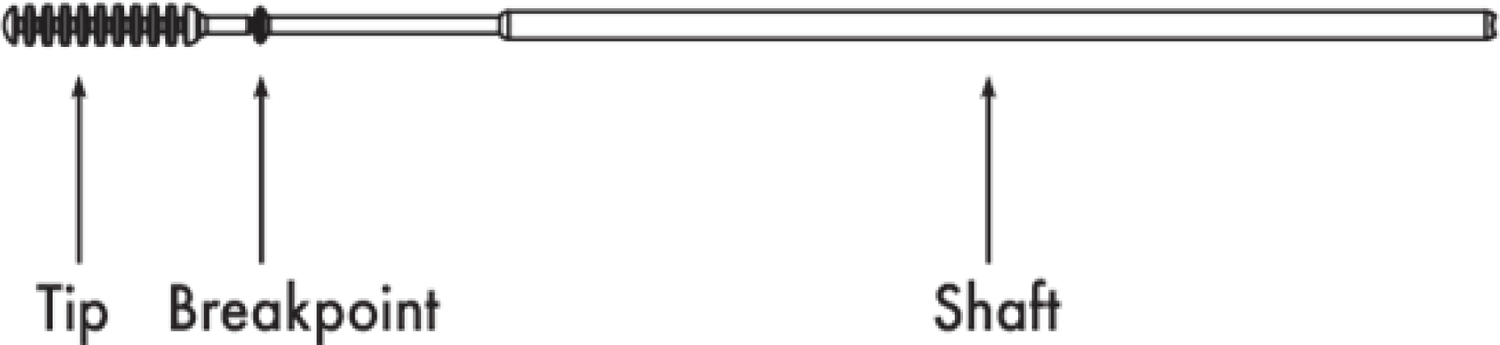
Developing a automation compatible swab. A major bottleneck is the sample processing is the manual process of opening tubes and removing the swab. Not only is this manual but also a source of both cross contamination and biohazardous exposure. To limit this, we designed a 3D printed swab, in conjunction with Applied Medical Company where the breakpoint was engineered to break close to the swab edge such that it would not interfere with our automated pipette machinery.

**Figure S14.**
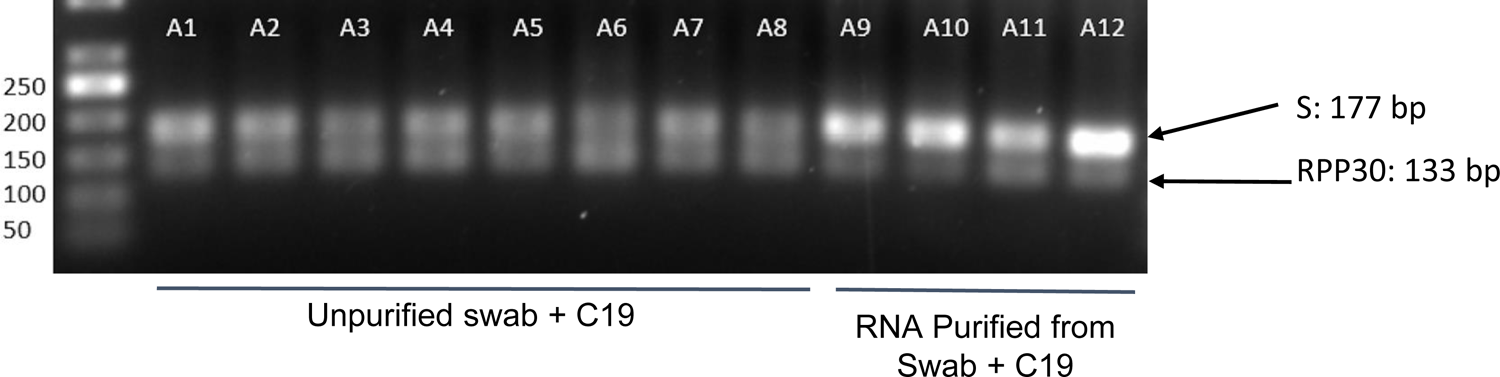
PCR inhibition has significant effect on amplification products. A) 2% Agarose gene was run for a subset of wells from our Rt-PCR reactions. We observe RT-PCR inhibition from swabs in unpurified lysate (A1-A8) as compared to purified RNA (A9-A12). We observe two bands in this subset of wells representing 2 amplicons for the S or S spike (177bp) and RPP30 (133 bp) primer pairs.

**Figure S15.**
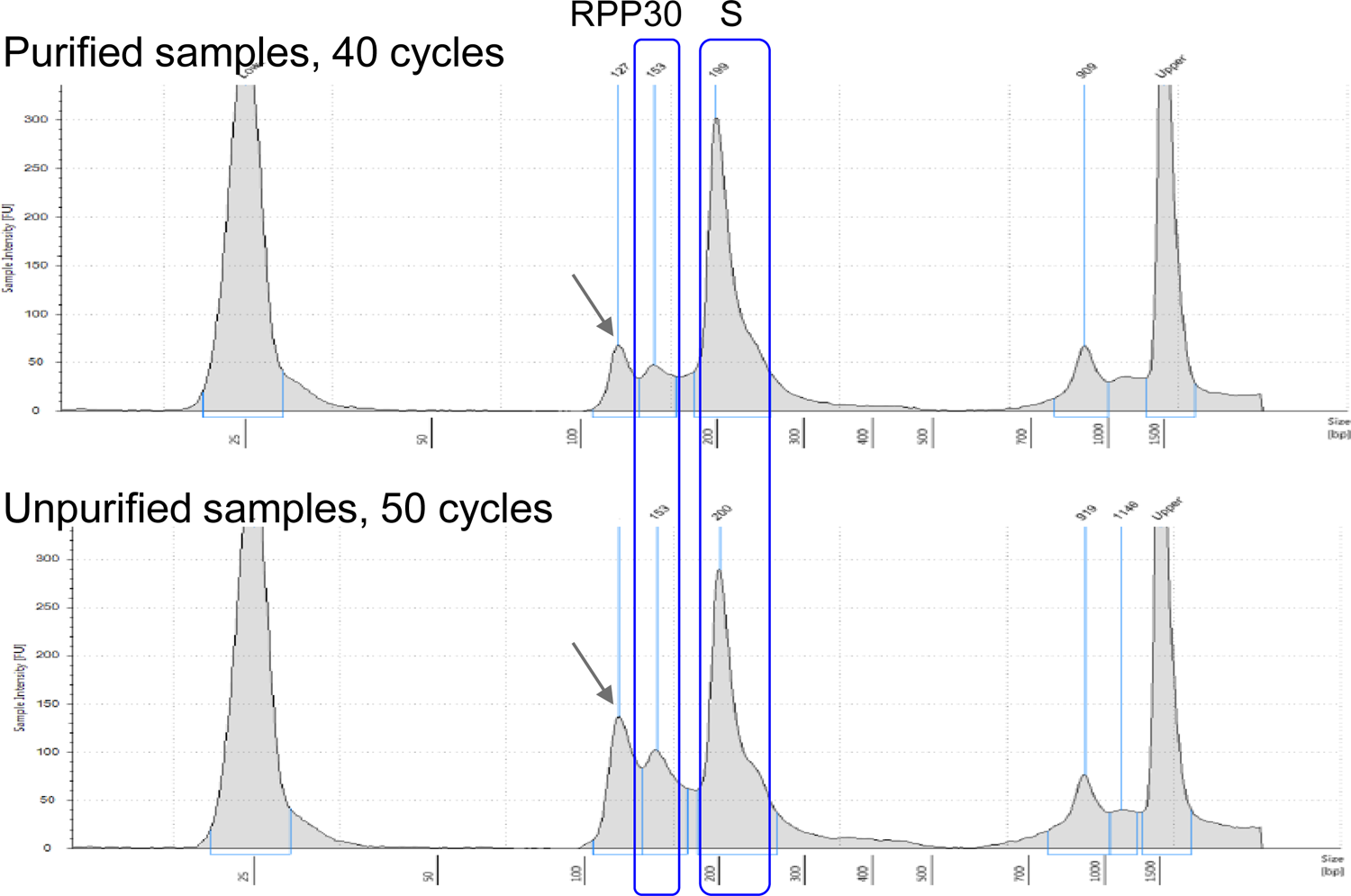
Tapestation Increasing the number of PCR cycles and working with unpurified or inhibitory samples types (eg. Saliva) was seen to increase the size of a nonspecific peak in our library preparation. Representative result from Agilent TapeStation for our purified amplicon libraries. We observe a nonspecific peak slightly above 100bp (arrow) in both library traces, but this peak increases in size with unpurified samples and an increased number of PCR cycles. While we have not confirmed the identity of this peak, we believe this peak may be the result of adapter dimers or unsequenceable PCR artifacts. Importantly, we observe that an increase in the size of this nonspecific peak leads to inaccurate library quantification. Therefore, in order to optimize cluster density on Illumina sequencers, we suggest quantifying the loading concentration of the final library based on the proportion of the desired peaks (RPP30 and S).

**Figure S16.**
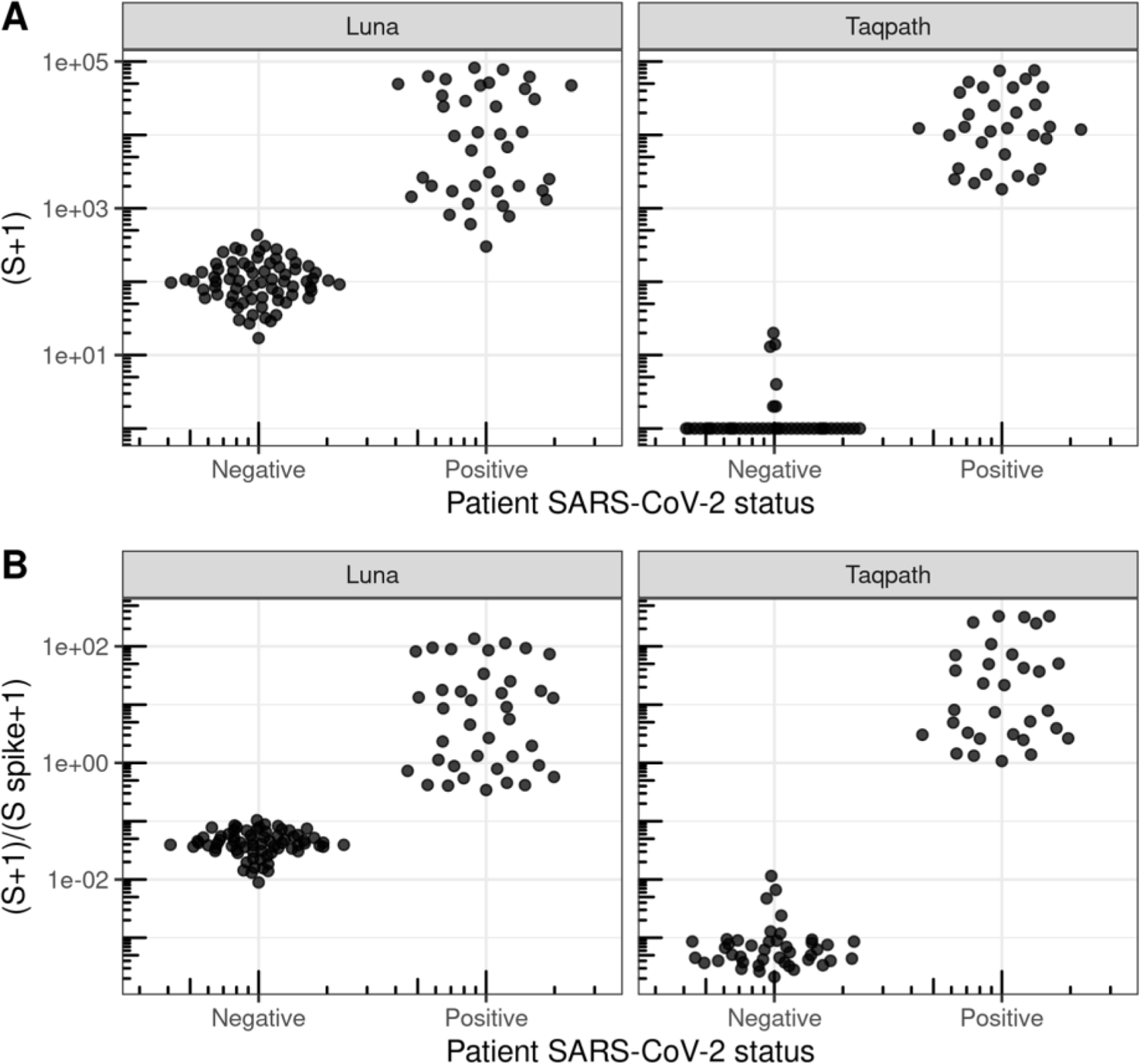
TaqPath decreases the number of S reads in SARS-CoV2-negative samples relative to NEB Luna. We compared Luna One Step RT-PCR Mix (New England Biosciences) to TaqPath™ 1-Step RT-qPCR Master Mix (Thermofisher Scientific). It is likely that the presence of UNG in the TaqPath Mastermix significantly reduced the number of S reads in the SARS-CoV-2-negative samples allowing us to more accurately distinguish SARS-CoV-2-positive and SARS-CoV-2-negative samples.

**Figure S17.**
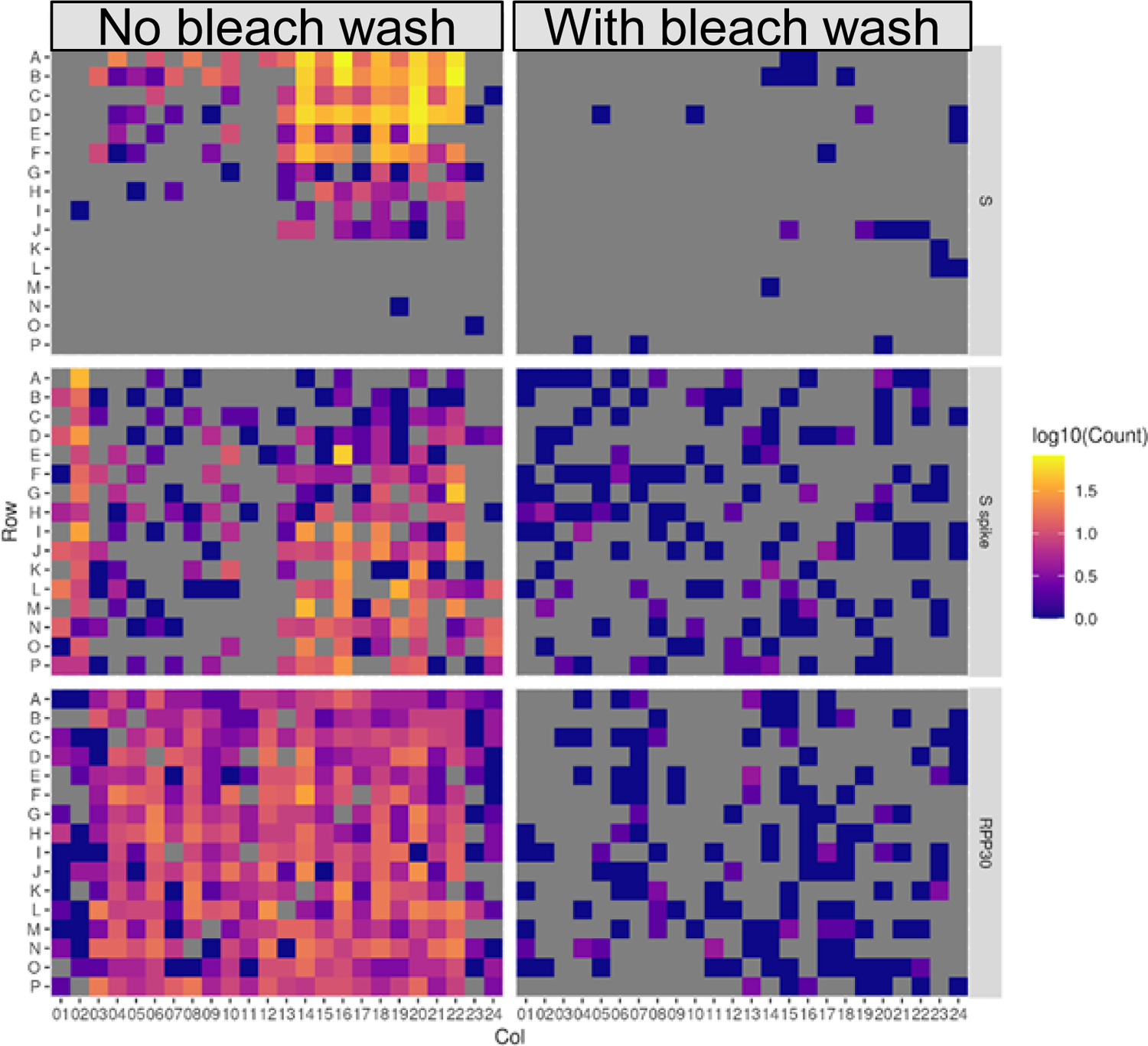
Carryover contamination from template line in a MiSeq contributes to cross contamination. In this experiment we did RT-PCR on four 384-well plates but only pooled three plates. On the left are observed counts of each of the amplicons for each sample for the 384-well plate not included in our run (but for which the indices were used in the previous run). Amplicon reads for indices used in the previous run are present at a low level (0-150 reads). We then performed a bleach wash in addition to regular wash prior to the subsequent run. In this subsequent run, we pooled three different plates and left out the fourth 384 well plate. On the right are observed counts of each of the amplicons for sample indices corresponding to the left-out plate (again, for which the indices were used in the previous run). We observe a remarkable decrease in the amount of carryover contamination, where carryover reads are <10 per sample.

**Figure S18.**
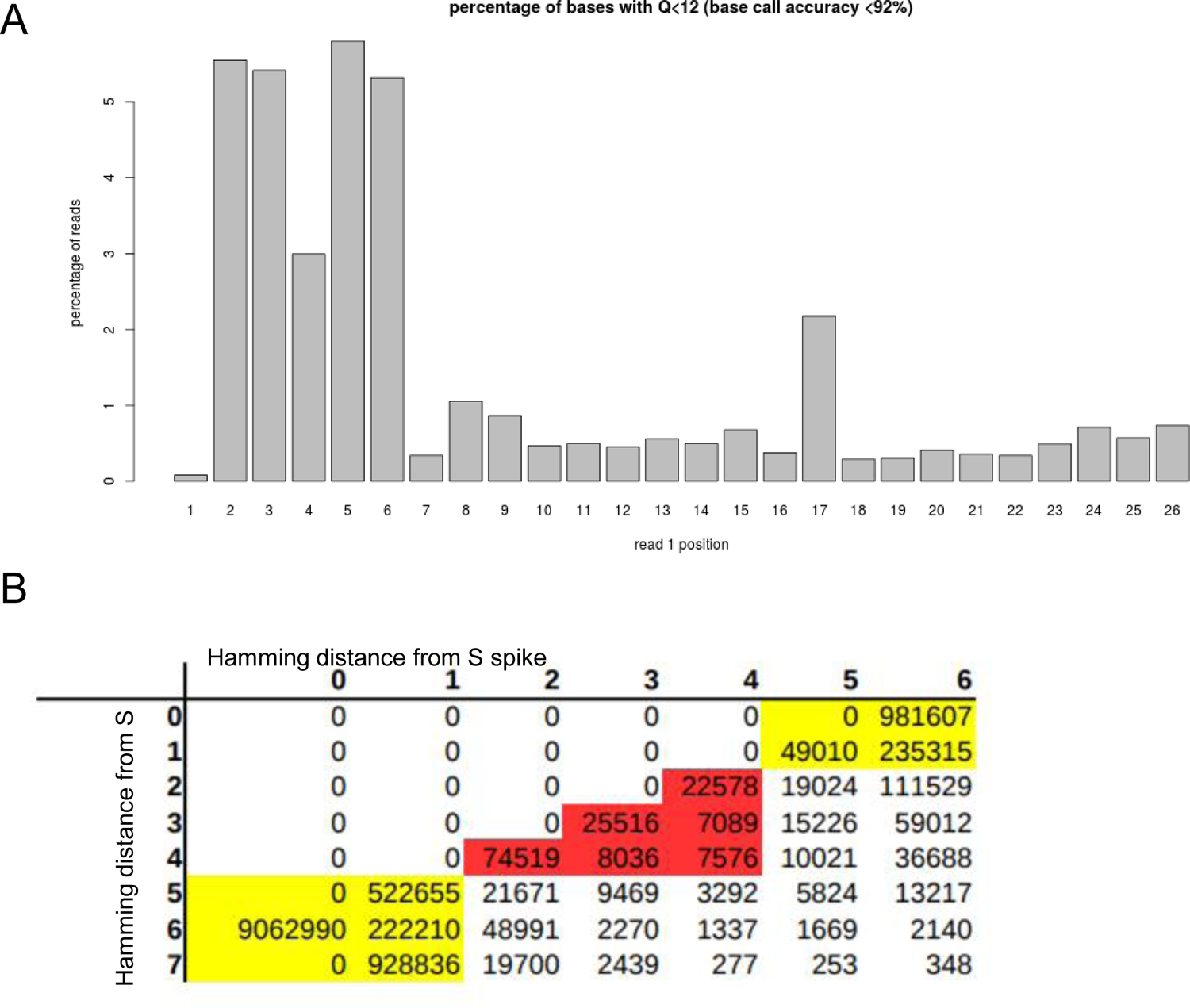
Sequencing errors in amplicon read and potential amplicon mis-assignment. In experiment v18 we loaded less PhiX than usual (11%) and the overall quality of read1 was lower. Trends noticed here persist in other runs but this run more clearly highlights issues that can occur due to sequencing errors and overly tolerant error-correction. A) The percentage of reads with base quality scores less than 12 for each position in read 1. Note that the first 6 bases of read1 distinguish S from S spike and have the highest percentage of low quality base calls. B) The hamming distance between each read1 sequence and either the expected S sequence (rows) or S spike sequence (columns), In yellow are perfect match and edit distance 1 sequences that can be clearly identified as S or S spike. In red are sequences with errors that may be mis-assigned (S spike assigned as S is most problematic for this assay.)

**Figure S19.**
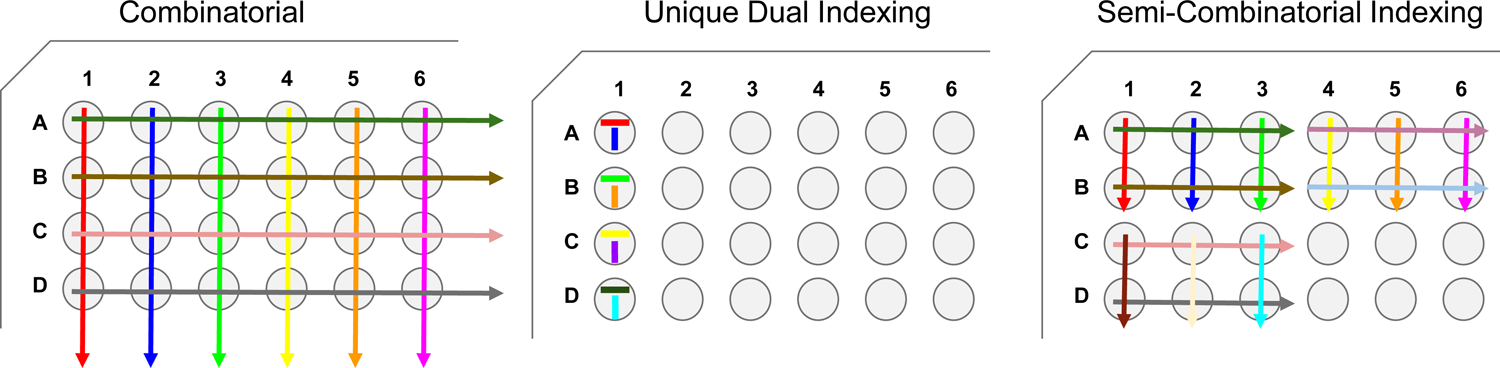
Visualization of different indexing strategies. Here i5 indices are depicted as horizontal lines, i7 indices are depicted as vertical lines, and colors represent unique indices. In combinatorial (or fully-combinatorial) indexing, the i5 and i7 indices are combined to make unique combinations, but each i5 and i7 index may be used multiple times within a plate, and all possible i5 and i7. For unique dual indexing, each i5 and i7 index are only used 1 time per plate. This requires many more oligos to be synthesized. For Semi-Combinatorial indexing, the combinations used are more limited, such that indices are only repeated for a subset of wells and many possible combinations are not used. In practice (not depicted here), we’ve used a design where the i7 index is unique but the i5 index can be repeated up to four times across a 384-well plate. For the majority of our SwabSeq development, we used either semi-combinatorial indexing (384×96) that allowed for 1536 combinations or samples to be run or unique dual indexing (384 UDI)

**Figure S20.**
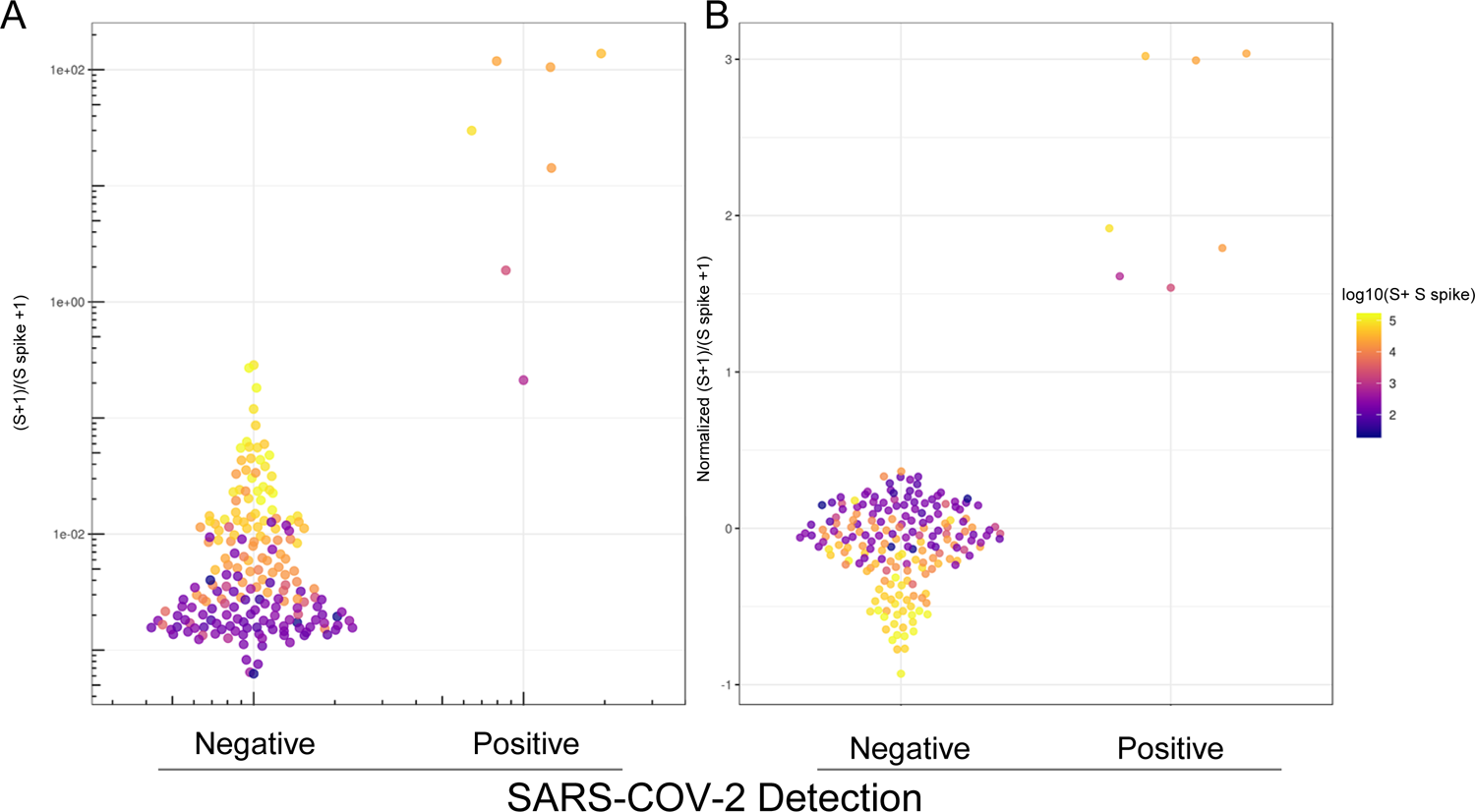
Computational correction for index mis-assignment using a mixed-model. To expand the number of samples we are capable of testing, we can use a combinatorial indexing strategy. In this experiment we used a single index on i5 to uniquely identify a plate and 96 i7 indices to identify wells. (A) The ratio of S to S spike (y-axis) is plotted for clinical samples based on whether Covid was detected by RT-qPCR (x-axis). SARS-CoV-2 positive samples were filtered to have Ct<32. The effects of index mis-assignment across plates can be observed as i7 indices that have high a sum of S and S spike across all samples that share the same i7 barcode across plates (colors). (B) Best linear unbiased predictor residuals are plotted (y-axis) for data in A, after computational correction of the log10(S+1/S spike+1) ratio by treating the identity of the i7 barcode as a random effect.

**Figure S21.**
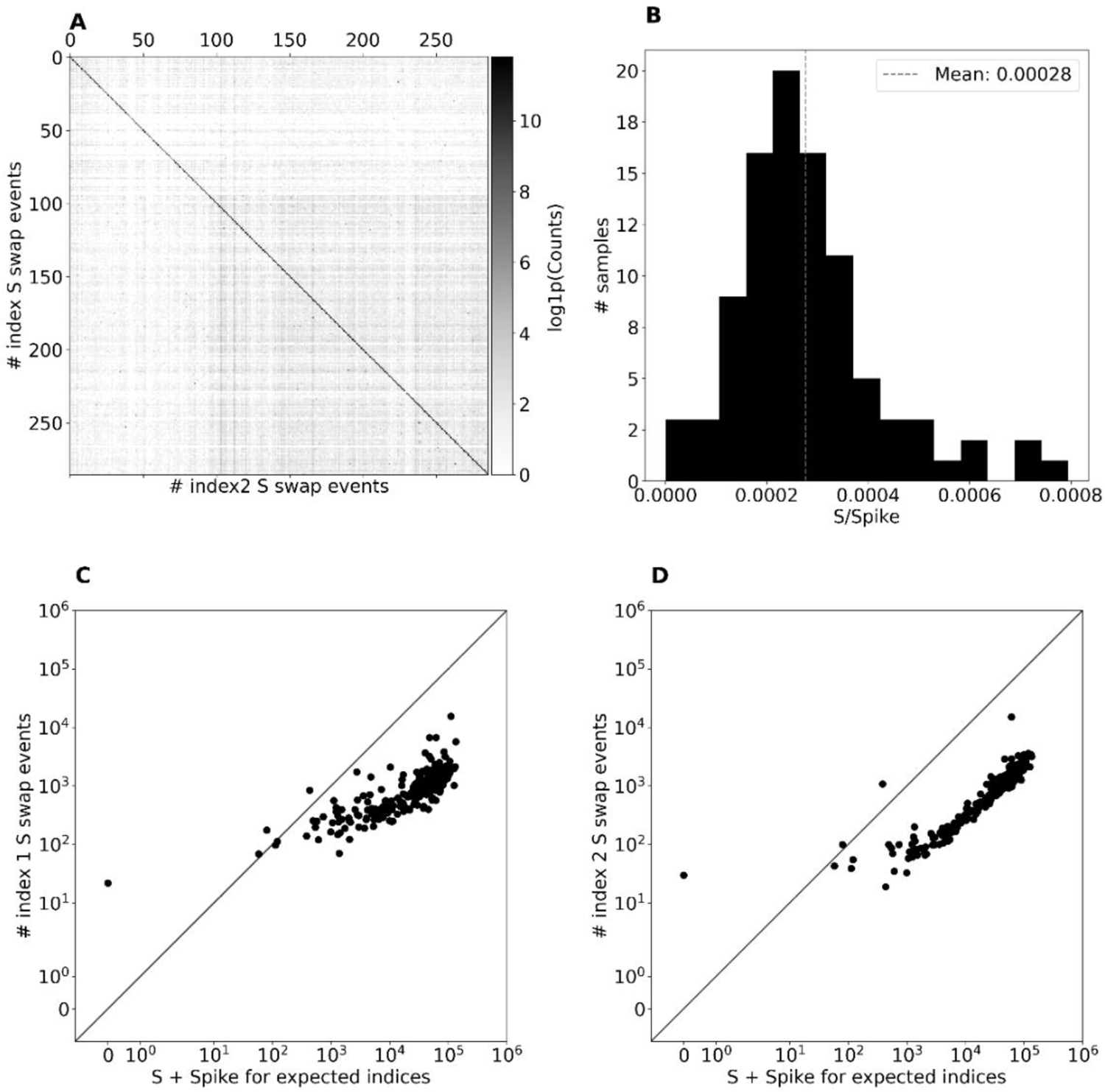
Quantifying the role of index mis-assignment as a source of noise in the S reads. A) A matching matrix for the viral S + S spike count for each pair of i5 and i7 index pairs from run v19 that used a unique dual index design. The index pairs along the diagonal correspond to expected index pairs for samples present in the experiment (expected matching indices) and the index pairs off of the diagonal correspond to index mis-assignment events. B) The distribution of ratios of viral S counts to Spike counts for samples with known zero amount of viral RNA. The mean ratio is 0.00028. C) The number of i7 mis-assignment events vs the number of viral S + S Spike counts for each sample. D) The number of i5 mis-assignment events vs the number of viral S + S Spike counts for each sample.

**Figure S22.**
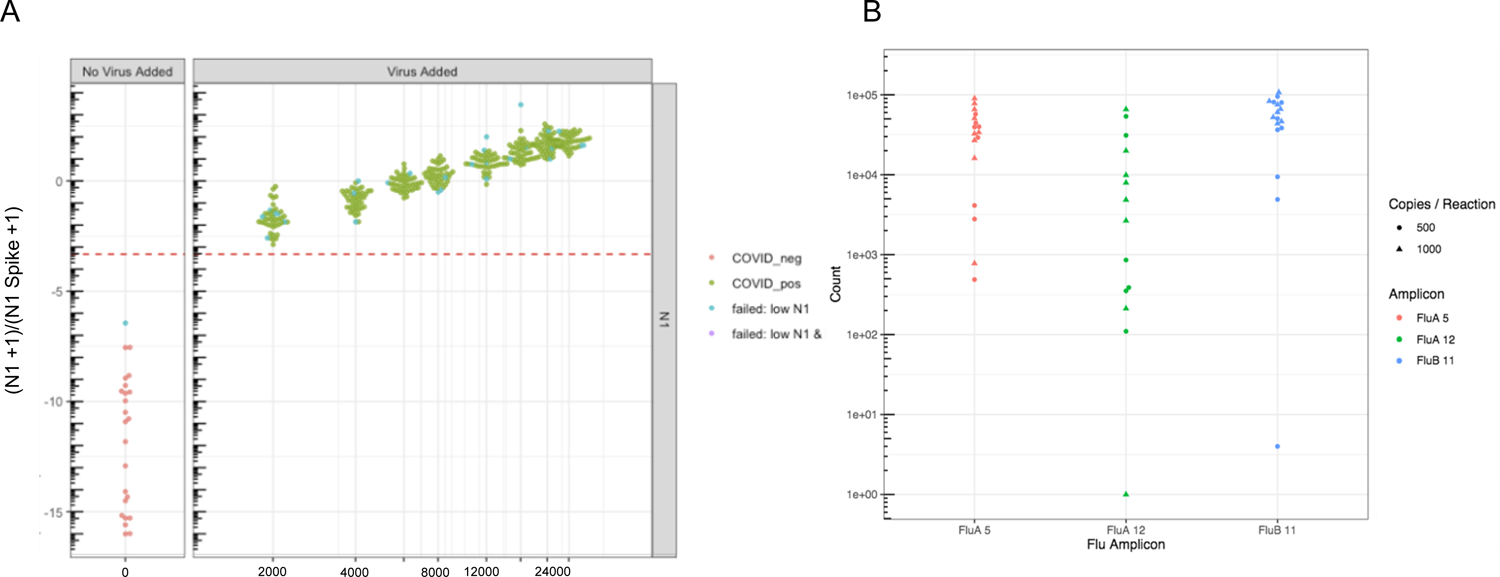
Extension of the SwabSeq Assay. A) We developed and tested to multiplex with additional SARS-CoV-2 amplicons. Here we demonstrate the N1 Amplicon LOD in saliva is around 2000 copies per mL. B) Testing of the three flu amplicons demonstrates that we are able to detect by sequencing samples with 500-1000 copies / reaction in PBS.

## REFERENCES

1. Furukawa NW, Brooks JT, Sobel J. Evidence Supporting Transmission of Severe Acute Respiratory Syndrome Coronavirus 2 While Presymptomatic or Asymptomatic. Emerg Infect Dis 2020; 26. doi:10.3201/eid2607.201595.

2. Lavezzo E, Franchin E, Ciavarella C, Cuomo-Dannenburg G, Barzon L, Del Vecchio C et al. Suppression of a SARS-CoV-2 outbreak in the Italian municipality of Vo’. Nature 2020. doi:10.1038/s41586-020-2488-1.

3. Peiris JSM, Yuen KY, Osterhaus ADME, Stöhr K. The severe acute respiratory syndrome. N Engl J Med 2003; 349: 2431–2441.

4. Cheng C, Wong W-M, Tsang KW. Perception of benefits and costs during SARS outbreak: An 18-month prospective study. J Consult Clin Psychol 2006; 74: 870–879.

5. Gandhi M, Yokoe DS, Havlir DV. Asymptomatic Transmission, the Achilles’ Heel of Current Strategies to Control Covid-19. N. Engl. J. Med. 2020; 382: 2158–2160.

6. Kimball A, Hatfield KM, Arons M, James A, Taylor J, Spicer K et al. Asymptomatic and Presymptomatic SARS-CoV-2 Infections in Residents of a Long-Term Care Skilled Nursing Facility - King County, Washington, March 2020. *MMWR Morb Mortal Wkly Rep* 2020; **69**: 377–381.

7. Fernandez M, Mervosh S. Texas Pauses Reopening as Virus Cases Soar Across the South and West. The New York Times. 2020.https://www.nytimes.com/2020/06/25/us/texas-coronavirus-cases-reopening.html (accessed 25 Jun2020).

8. News Division. HHS Details Multiple COVID-19 Testing Statistics as National Test. https://www.hhs.gov/about/news/2020/07/31/hhs-details-multiple-covid-19-testing-statistics-as-national-test-volume-surges.html (accessed 3 Aug2020).

9. CMS Increases Medicare Payment for High-Production Coronavirus Lab Tests. https://www.cms.gov/newsroom/press-releases/cms-increases-medicare-payment-high-production-coronavirus-lab-tests-0 (accessed 3 Aug2020).

10. Kliff S. Most Coronavirus Tests Cost About 100. Why Did One Cost 2,315? The New York Times. 2020. https://www.nytimes.com/2020/06/16/upshot/coronavirus-test-cost-varies-widely.html (accessed 25 Jun2020).

11. Pollitz K. Free Coronavirus Testing for Privately Insured Patients? Kasier Family Foundation. 2020.https://www.kff.org/coronavirus-policy-watch/free-coronavirus-testing-for-privately-insured-patients/ (accessed 25 Jun2020).

12. Larremore DB, Wilder B, Lester E, Shehata S, Burke JM, Hay JA et al. Surveillance testing of SARS-CoV-2. Infectious Diseases (except HIV/AIDS). 2020. doi:10.1101/2020.06.22.20136309.

13. Paltiel AD, David Paltiel A, Zheng A, Walensky RP. Assessment of SARS-CoV-2 Screening Strategies to Permit the Safe Reopening of College Campuses in the United States. JAMA Network Open. 2020; 3: e2016818.

14. Eric M Jones, Aaron R Cooper, Joshua S Bloom, Nathan B. Lubock, Scott W. Simpkins, Molly Gasperini, Sriram Kosuri. Octant SwabSeq Testing. 2020.https://www.notion.so/Octant-SwabSeq-Testing-9eb80e793d7e46348038aa80a5a901fd.

15. Jones EM, Jajoo R, Cancilla D, Lubock NB, Wang J, Satyadi M et al. A Scalable, Multiplexed Assay for Decoding GPCR-Ligand Interactions with RNA Sequencing. Cell Syst 2019; 8: 254–260.e6.

16. Schmid-Burgk JL, Li D, Feldman D, Słabicki M, Borrajo J, Strecker J, et al. LAMP-Seq: Population-Scale COVID-19 Diagnostics Using a Compressed Barcode Space. bioRxiv. 2020;: 2020.04.06.025635.

17. COVIDSeq Test (RUO and PEO Versions). https://www.illumina.com/products/by-type/clinical-research-products/covidseq.html (accessed 3 Aug2020).

18. rapidmicrobiology Bioinnovation’s DxSeq^TM^ Sequences Filoviruses. https://www.rapidmicrobiology.com/news/bioinnovations-dxseq-sequences-filoviruses (accessed 3 Aug2020).

19. Booeshaghi AS, Lubock NB, Cooper AR, Simpkins SW, Bloom JS, Gehring J et al. Fast and accurate diagnostics from highly multiplexed sequencing assays. Health Informatics. 2020. doi:10.1101/2020.05.13.20100131.

20. Relich RF. PREPARATION OF VIRAL TRANSPORT MEDIUM. 2020 https://www.cdc.gov/coronavirus/2019-ncov/downloads/Viral-Transport-Medium.pdf.

21. CDC. Information for Laboratories about Coronavirus (COVID-19). Centers for Disease Control and Prevention. 2020.https://www.cdc.gov/coronavirus/2019-ncov/lab/rt-pcr-panel-primer-probes.html (accessed 6 Jul2020).

22. Ranoa DRE, Holland RL, Alnaji FG, Green KJ, Wang L, Brooke CB, et al. Saliva-Based Molecular Testing for SARS-CoV-2 that Bypasses RNA Extraction. bioRxiv. 2020;: 2020.06.18.159434.

23. COVID-19 Testing at Broad. https://covid-19-test-info.broadinstitute.org/ (accessed 7 Jul2020).

24. iSWAB Rack Format - Mawi DNA Technologies. Mawi DNA Technologies. https://mawidna.com/our-products/iswab-rack-format/ (accessed 7 Jul2020).

25. Covid T. Multiplex Diagnostic Solution| Thermo Fisher Scientific-US. 19AD.

26. Clark TW, Brendish NJ, Poole S, Naidu VV, Mansbridge C, Norton N et al. Diagnostic accuracy of the FebriDx host response point-of-care test in patients hospitalised with suspected COVID-19. J Infect 2020. doi:10.1016/j.jinf.2020.06.051.

27. Yujian L, Bo L. A normalized Levenshtein distance metric. IEEE Trans Pattern Anal Mach Intell 2007; 29: 1091–1095.

28. Untergasser A, Cutcutache I, Koressaar T, Ye J, Faircloth BC, Remm M et al. Primer3--new capabilities and interfaces. Nucleic Acids Res 2012; 40: e115.

29. Quail MA, Swerdlow H, Turner DJ. Improved protocols for the illumina genome analyzer sequencing system. Curr Protoc Hum Genet 2009; **Chapter 18**: Unit 18.2.

30. Morgan M, Anders S, Lawrence M, Aboyoun P, Pagès H, Gentleman R. ShortRead: a bioconductor package for input, quality assessment and exploration of high-throughput sequence data. Bioinformatics 2009; 25: 2607–2608.

31. Van der Loo MPJ. The stringdist package for approximate string matching. R J 2014; 6: 111–122.

32. Booeshaghi AS, Lubock NB, Cooper AR, Simpkins SW, Bloom JS, Gehring J et al. Reliable and accurate diagnostics from highly multiplexed sequencing assays. Sci Rep 2020; 10: 21759.

33. Melsted P, Sina Booeshaghi A, Gao F, Beltrame E, Lu L, Hjorleifsson KE, et al. Modular and efficient pre-processing of single-cell RNA-seq. bioRxiv. 2019;: 673285.

34. Furtado LV, Gong J, Huard T, Jeng L, Joseph L, Kim A et al. The 2013 AMP Clinical Practice Committee consisted of Matthew J. Bankowski, Milena Cankovic, Jennifer Dunlap. https://www.amp.org/AMP/assets/File/resources/201503032014AssayValidationWhitePape r.pdf?pass=24.

35. Nelson MC, Morrison HG, Benjamino J, Grim SL, Graf J. Analysis, optimization and verification of Illumina-generated 16S rRNA gene amplicon surveys. PLoS One 2014; 9: e94249.

36. Valk T van der, van der Valk T, Vezzi F, Ormestad M, Dalén L, Guschanski K. Index hopping on the Illumina HiseqX platform and its consequences for ancient DNA studies. Molecular Ecology Resources. 2019. doi:10.1111/1755-0998.13009.

37. MacConaill LE, Burns RT, Nag A, Coleman HA, Slevin MK, Giorda K et al. Unique, dual-indexed sequencing adapters with UMIs effectively eliminate index cross-talk and significantly improve sensitivity of massively parallel sequencing. BMC Genomics 2018; 19: 30.

