## Supplementary material for "Swab-Seq: A high-throughput platform for massively scaled up SARS-CoV-2 testing": Optimized Protocols

### 1. **PURPOSE**

SwabSeq technology presents a unique approach to viral testing that eliminates many of the current bottlenecks that limit clinical testing and importantly can rapidly scale to 10,000 samples per day with simple automation and many of the machines and semi-automation that are standard in research laboratories. In this operating procedure, we provide the list of reagents and protocols and in addition we outline the methods for using SwabSeq with alternative specimen types such as extraction free saliva and extraction free nasal swabs.

### 2. **MATERIALS**

**NOTE: Do not use any reagent beyond the expiration date**

Table 1. List of reagents.

| **Reagent** | **Storage Temperature** | **Preparation Details (note here or refer to procedure)** |
| --- | --- | --- |
| S Standard | -80°C | See Section C, page 6 |
| Primers | -20°C | See Section B, page 3 |
| magMAX Viral/Pathogen Nucleic Acid Isolation Kit | 4°C-8°C and RT | See Section D, Page 9 |
| TBE Buffer (Tris-borate-EDTA) (10X) | 15°C -30°C | See Section E, page 10 |
| Nuclease Free Water | 15°C -30°C | See Section F, page 10 |
| Tween-20 | 15°C -30°C | See Section E, page 22 |
| TaqPath™ 1-Step RT-qPCR Master Mix (Fisher Scientific, A15300) | -20°C | See Section G, page 12 |
| Ampure XP Beads (Fisher Scientific, A63880) | 4°C-8°C | See Section H, page 14 |
| Qubit™ RNA HS Assay Kit (ThermoFisher, Q32855) | 4°C-8°C and RT | See Section H, page 14 |
| PhiX Control v3 (Illumina, FC-110-3001) | 4°C-8°C and RT | See Section H, page 14 |
| MiSeq Reagent Kit v3 (150-cycle, Catalog # MS-102-3001) | -20°C | See Section I, page 15 |
| NextSeq 500/550 High Output Kit v2 (150 cycles, #20024907) | -20°C | See Section I, page 15 |

Table 2. Acceptable specimen, storage and handling.

| **Acceptable Specimens** | **Storage Temperature** | **Storage Time** |
| --- | --- | --- |
| Nasal Swabs | 4°C, -20°C | 72 hours, 1 week |
| Saliva | 4°C, -20°C | 72 hours, 1 week |

#### 2.1. Reagents and equipment

General

Capit-All (Thermofisher Scientific)

IntelliXcap Capper Decapper (Brooks)

Pipette Tips LQR LTS 20μL FL 960/10 (Rainin, 17014400 for Bench Smart)

12.5 μl GRIPTIP Sterile, Filter, LONG, 5 XYZ Racks of 384 Tips, Low Retention (Integra , 6505)

384-WELL HARDSHELL PLATE CLEAR (#20 PCS PK 4483285 Fisher)

TaqPath™ 1-Step RT-qPCR Master Mix (Fisher Scientific, A15300)

TE Buffer, Tris-EDTA, 1X Solution, pH 7.4 (Fisher BP24761)

Tween-20

EtOH

NUCLEASE-FREE WATER (Thermofisher, AM9937)

96-well plates (qRT-PCR)

Normal Saline

10X TBE 1M Tris-HCl pH8.0, 900mM boric acid, and 10mM EDTA(10x Buffer, ThermoFisher)

Synthetic S Standard Construction

### HiScribe™ T7 High Yield RNA Synthesis Kit (New England Biolabs, E2040S)

### DNAse, RNAse Free (NEB, M0303S)

### RNA Clean & Concentrator-5 (Zymo, [R1013](https://www.zymoresearch.com/collections/rna-clean-concentrator-kits-rcc/products/rna-clean-concentrator-5))

Post-PCR Pooling and Purification

DynaMag-2 Magnet (Thermofisher Scientific, 12321D)

Ampure XP Beads (Fisher Scientific, A63880)

1.7 mL Eppendorf Lo-Bind Tubes (Fisher Scientific, 13-698-791)

150 mL reservoir, sterile, bulk, automation friendly, polystyrene (Integra, 6318)

Qubit™ RNA HS Assay Kit (ThermoFisher, Q32855)

Qubit™ Assay Tubes (Thermofisher Scientific, Q32856)

Qubit™ RNA BR Assay Kit (ThermoFisher, Q10211)

Sequencing Materials and Reagents

PhiX Control v3 (Illumina, FC-110-3001)

Illumina Free Adapter Blocking Reagent (48 reactions, # 20024145)

MiSeq Reagent Kit v3 (150-cycle, Catalog # MS-102-3001)

NextSeq 500/550 High Output Kit v2 (150 cycles, #20024907)

#### 2.2. Primers

**Indexed Primers**

Custom Primer sets were designed to amplify the S gene in the SARS-CoV2 genome and the RPP30 gene in the human genome. The S gene amplification indicates the presence of SARS-CoV2 RNA genome within the specimen. The RPP30 gene demonstrates adequate sample collection. Each primer pair (i5 and i7, designated as F and R) has an 10-basepair random barcode adapted to the primer that we have designed to be unique to each i7 barcode which represents the plate-well for that barcode. The i5 barcode can be repeated over the plate and is sometimes referred to as the “plate barcode”. The combinatorial indexing strategy limits the number of unique primers that need to be purchased. An alternative, but more expensive strategy, would be to purchase unique-dual indices (UDI) which are designed to be a unique pair in each well.

**S and RPP30 Primers**, Custom Primer Design is attached as a separate document “UCLA_SwabSeq_barcodedPrimers_V4”

Ordered from IDT

100nM, 200uM concentration, Standard Purification

**Final Concentration for Working Primers (10x):**

4 μM for S

0.5 μM for RPP30

**Custom Sequencing Primers**

These primers are for custom sequencing approaches on the Illumina based flow cell.

Ordered from Integrated DNA Technologies

100nM synthesis scale, concentration is 100uM

Table 3. Primers used for specific amplification of S gene and human control RPP30.

| **S_SARS-CoV-2** |  | **Tm** |
| --- | --- | --- |
| Read_1 | gctggtgctgcagcttattatgtgggt | 63 |
| i7_seq | agatgctgtagactgtgcacttgaccct | 63 |
| i5_seq | acccacataataagctgcagcaccagc | 63 |
| **RPP30** |  |  |
| Read_1 | gagcggctgtctccacaagtccg | 63.5 |
| i7_seq | acccgctcgcaggtccaaatct | 62.6 |
| i5_seq | cggacttgtggagacagccgctc | 63.5 |

### 3. **METHOD**

#### 3.1. Preparing Primer Plates

All plates will be pre-stamped with a mix of indexed primers that are 10x concentration (4 μM for SFor and SRev; 0.5 μM for RPP30For and RPP30Rev). For a 20 μL reaction, we will place 2uL into each well of the primer plate. These were ordered in bulk from IDT.

1) Spin down master plates at 2000xg for 1 minute to ensure that all frozen ice is at the bottom of the well. This prevents cross contamination of indexed primers when removing the foil lid.
2) Set up Integra work station:
   a) Scan or write down Barcode Labels in the Primer Plate Notebook
   b) Label multiple 384-well primer plates with primer set name.
   c) Change the Setting on the Integra for multi-dispensing of 2uLs per plate x 6 plates
3) Remove seal from the master plates very carefully
4) Using the 384-well head for the Integra Viaflo, carefully pipet up primer plates and dispense 2uL into each plate.
5) Seal each plate, freeze in -20 until use.

Repeat with each 384-well Primer Plate Set.

An alternative approach is to purchase pre-stamped primer plates at the needed concentrations directly from the company synthesizing the plates. We have explored these options with a number of oligo synthesizing companies and have found that although it requires validation and resources for set up, it can drastically reduce the labor required to accurately pipet the primer plates.

#### 3.2. Construction and dilution of *in vitro* S RNA standard

**Purpose:** We use an *in vitro* S RNA standard as a control in each of our wells. The sequence is meant to mimic the actual SARS CoV-2 amplicon (same amplicon structure except for a 6 nt unique stretch to distinguish). The use of this synthetic standard has two important advantages. This can be done in large batches every 6-months to 1-year or as needed depending on the rate of usage.

For this in vitro standard, sequencing requires the addition of high percentage of PhiX in order to assist with cluster generation. To simplify our process, we currently use the diversified standard (section 3.4) that was pre-synthesized for our current testing needs.

1. This serves as an in-well positive control for the S primers. Even in a sample that has no SARS-CoV-2, we can observe amplification with primers, thereby ensuring that a negative result is due to lack of virus, and not due to technical issues with the primers.
2. A second benefit is that we can use the ratio between S/S standard in our analysis pipeline. This has the effect of demonstrating that small changes in experimental conditions due to differences in samples, noise and amplification biases are not driving our results.

**RT-PCR primers for standard constriction using *In vitro* Transcription (adding on T7 promoter)**

Table 4. Primers to construct template for the S Synthetic standard.

| S_FP | TAATACGACTCACTATAGggctggtgctgcagcttattatgtgggtATAGAAcaacctaggacttttctattaa |
| --- | --- |
| S_RP | aacgtacactttgtttctgagagagg |

1. Perform RT-PCR in a 96-Well thermocycler using the primers above and gRNA of SARS-CoV-2.
2. Run on an Agarose gel and ensure specific products at ~130 bp
3. Purify DNA using Ampure Beads, use a ratio of 1.8 ratio of beads: sample volume.
4. Vortex and let sit for 5 minutes at room temperature.
5. Use magnet to collect beads for 1 minute.
6. Remove liquid and wash beads twice with 500 ul of freshly made 70% EtOH.
7. Elute in 100uL of 0.1X Qiagen EB buffer.
8. Use a magnet to collect beads for 1 minute. Transfer 90 uL to new eppendorf tube.
9. Quantify samples using Qubit DNA BR Kit.

Store DNA overnight or at -20 for long term storage if not immediately proceeding to IVT reaction.

10. Thaw the necessary kit components from the NEB HiScribe Kit. Mix and pulse-spin in microfuge to collect solutions to bottom of tubes. Keep on ice.
11. Prepare MasterMix for In Vitro Translation Reaction:
    a. If you are planning to run many reactions, it is convenient to prepare a master mix by combining equal volumes of the 10X reaction buffer and four ribonucleotide (NTP) solutions
    b. Using this mastermix (see below Table 1) we made a 4 reactions.
    c. Split into 4 tubes with 20 μl each.
    d. Vortex, pulse spin, but at 37˚C overnight in Thermocycler machine.

Table 5. MasterMix for IVT to make Synthetic Standard S

| **Component** | **per reaction** | **Property** | **4 reactions (μl)** |
| --- | --- | --- | --- |
| Nuclease-free Water | 2.5 μl |  | 10 |
| 10X Reaction Buffer | 1.5 μl | 0.75X final | 6 |
| NTP | 1.5 μl each | 7.5 mM each final (4µL total) | 6 |
| Template DNA | 13 μl | ~300-600ng template | 52 |
| T7 RNA Polymerase Mix | 1.5 μl |  | 6 |
| Total reaction volume | 20 μl |  | 80 |

**Post-IVT Purification**

1. After IVT, we DNAse treated the reactions by adding 1uL of DNase (NEB, M0303S) to each reaction
2. Incubate reactions at 37°C for 10 minutes.
3. Add 1 µl of 0.5 M EDTA (to a final concentration of 5 mM).
4. Heat inactivate at 75°C for 10 minutes.
5. RNA is purified using the [Zymo RNA clean and concentrator column](https://www.zymoresearch.com/collections/rna-clean-concentrator-kits-rcc).
6. RNA is quantified with an [Agilent TapeStation](https://www.agilent.com/en/product/automated-electrophoresis/tapestation-systems/tapestation-rna-screentape-reagents/rna-screentape-analysis-228268) or Qubit using the RNA BR Kit.

#### 3.3.  *In vitro* S standard dilution protocol

The Synthetic S standard is an internal well control for the S primer pair. This standard is placed directly in the master mix and diluted to the same copy number in every well. Therefore, even for samples in which there is no SARS-CoV-2 virus present, we have an internal control demonstrating that the reaction conditions were sufficient for amplification with the S primer pair. The key for the synthetic S standard is to include it at a concentration of 100-1000 copies per reaction. Therefore, quantitation of the synthetic S standard is key to this experiment; if the quantitation is off by an order of magnitude, the synthetic standard can overwhelm signal seen in the experiment. Our current working protocol uses 250 copies per reaction.

Due to the sensitivity of the standard dilution quantitation, we suggest making small aliquots that are thawed per run every month and performing RT-qPCR to quantify the copies/reaction. Aliquots are then created from this batch and stored at -80०C to minimize freeze thaws.

**Protocol for Creating Standard Dilutions:**

This should be done fresh every month to prevent degradation of the standard dilutions. Each month, enough aliquots should be created to last the estimated number of runs for the month.

1) Clean Biosafety Hood with 10% bleach and treat bench with UV light for 15 minutes
2) Treat pipettes, 0.1% Tween-20 in water with UV light for 15 minutes
3) Dilute and aliquot standard dilution into single use tubes:
   a) Measure the concentration of the S stock using HS RNA Qubit.
   b) Using the concentration from the previous step, calculate copies/uL using the equation found using the NEB Bio Calculator (nebiocalculator.neb.com). RNA length is 130 nt.
   c) Perform serial dilutions of 100-fold each using 0.1% Tween-20 in water until a concentration of ~10,000 copies/uL is reached. Save an aliquot of each dilution for quantification in step 5.
   d) Aliquot the final dilution into individual tubes containing 10uL each. These will be used to standard the master mix for each run of Swabseq.
4) *Perform a Qubit analysis on the first 100-fold dilution to validate.*
5) QC via RT-qPCR and validate copies/uL. Ct for 100 copies should come up between 33 and 34; for 1000 copies should come up around Ct of 30.

#### 3.4. Using the Diversified S RNA Standard

**Purpose:** Next-Generation Sequencing of a small number of amplicons requires the addition of PhiX standard to the library to provide sufficient diversity for cluster generation and short-read sequencing. We have developed a diversified version of the S RNA standard that provides sufficient library diversity without the need for PhiX. These can be purchased from an Synthego.

To prepare the diversified S RNA standard, prepare the 4 separate diversified S RNA standards as above. Combine equimolar concentrations of the 4 standards. Dilute according to the dilution protocol outlined above (Section 3.3).

Table 6: Diversified S RNA Standard Sequences

| S2_001 | GCTGGTGCTGCAGCTTATTATGTGGGTGTGTATCTCACGAAGCGACCCTTTGGAAAATATAATGAAAATGGAACCATTACAGATGCTGTAGACTGTGCACTTGACCCT |
| --- | --- |
| S2_002 | GCTGGTGCTGCAGCTTATTATGTGGGTCCTCGCTAGGACGTCGCTATgacgccAAAATATAATGAAAATGGAACCATTACAGATGCTGTAGACTGTGCACTTGACCCT |
| S2_003 | GCTGGTGCTGCAGCTTATTATGTGGGTAGCACGACTTGATCTAACTgacactaAAAATATAATGAAAATGGAACCATTACAGATGCTGTAGACTGTGCACTTGACCCT |
| S2_004 | GCTGGTGCTGCAGCTTATTATGTGGGTTAAGTAGGACTTCGATTggaTggaatAAAATATAATGAAAATGGAACCATTACAGATGCTGTAGACTGTGCACTTGACCCT |

#### 3.5. Samples testing on the SwabSeq Platform.

Swabseq can take in a variety of sample types. It performs very well with purified samples, however these are not as conducive to scale due to the labor-intensive RNA-purification process (Section 3.5.1). We have optimized our process with extraction free protocols for both nasal swabs into TE buffer (Section 3.5.2) and for saliva (Section 3.5.3)

#### 3.5.1. RNA-purified samples, Thermofisher Kingfisher platform

Nasal Swabs collected from into any of the traditional collection buffers (e.g., Aimes Buffer, Normal Saline or VTM) can be used for this protocol. Set up 96-well deep well plate for 94 samples. We use the same volumes, regardless of the type of collection media that is used. This process is performed at the UCLA Clinical Microbiology Laboratory in Brentwood Annex.

1) Fill plates with following liquids
   a. Wash Plate 1: 1mL Wash Buffer
   b. Wash Plate 2: 1mL 80% EtOH
   c. Elution Buffer Plate: 100 uL per well
2) Prepare Bead Binding Mix
   a. Prepare required amount on each day of use
   b. Vortex the total Nucleic Acid Magnetic Beads to ensure that the bead mixture is homogeneous
   c. Prepare the bead binding mix according to the table below

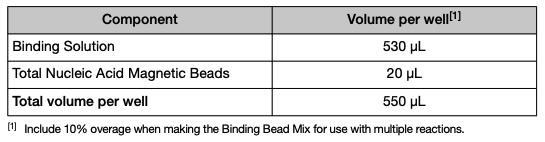

d. Mix well by inversion, store are room temperature until use

3) Digest with Proteinase K
   a. Add 1-uL of Proteinase K into each well of the Kingfisher Deepwell 96 plate
   b. Pipet 400uL of each patient sample into a 96-well format, leave PPC and NPC control well empty
   c. Add 400uL of Nuclease Free water to the negative control well
   d. Invert Binding Bead Mix 5 times gently to mix, then add 550 uL to each sample and negative control well
4) Seal the plate with the MicroAmp Clear Adhesive Film.
5) Load the King Fisher Flex Machine:
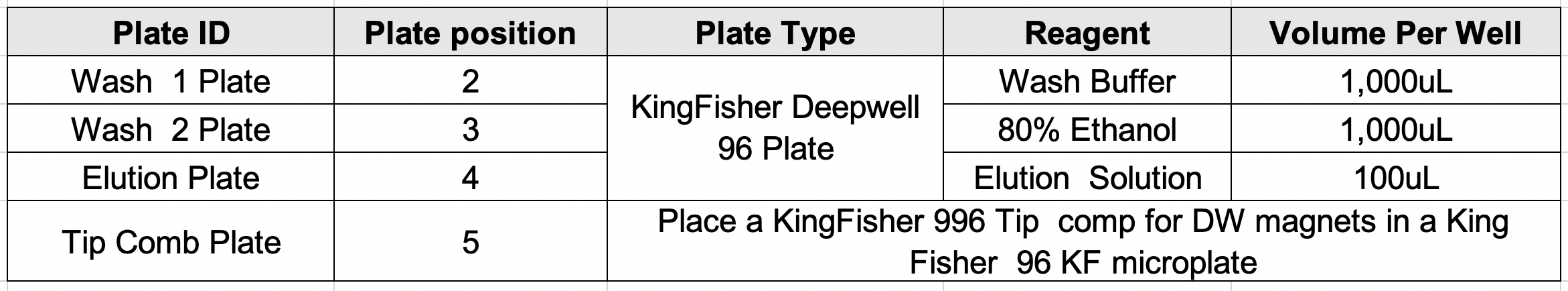

6) Select the MVP_2Wash_400_Flex on the KingFisher Flex Magnetic Particle Processor with the 96 Deep Well Head.
7) Start the run and load the prepared plates into position when prompted by the instrument
8) Remove the elution plate after the run is complete (~24 minutes later) and cover with Clear Adhesive Film.
9) Place elution plate on ice for use in RT-PCR until ready to load sample.
10) Pipet 7uL from deepwell sample plate into designated quadrant of 384 well plate using the Rainin Benchsmart 200uL.

#### 3.5.2. Saliva Specimen Processing

Saliva samples will be processed using extraction free methods. All samples will be received in the UCLA COVID19 testing lab in 650 Charles E Young Dr South, CHS Building for direct accessioning and processing.

1) Fresh Saliva is collected into a pre-barcoded tube by the user through a plastic funnel. All samples are stored and transported at 4°C.
2) Tubes with samples are placed in 96 well racks in a 95°C circulating water bath for 30 minutes.
3) Tubes are spun in 4°C centrifuge for 20 seconds at 200xg to collect saliva at bottom of tube.
4) Rack is placed in the automated decapper (Brooks Intellecap or ThermoFisher Capit-All) and the 96 tubes are uncapped.
5) Using the Benchsmart 200, we remove the topmost 200uL from the tubes at high speed to prevent overflow. Aspirate 40uL of saliva from just below the bottom of the tube and dispense back into the tube in increments of 5uL until there are no air bubbles at the bottoms of the tips. Dispense 5uL into the appropriate quadrant of the 384-well plate pre-loaded with primers and mastermix.
6) After each quadrant is dispensed into the 384-well plate, the 384 well plate is placed into the thermocycler and run with the RT-PCR reaction described below.

#### 3.5.3. Extraction Free Nasal Swab processing

Extraction Free Nasal Swab Specimens are NP, Mid-Nasal and Anterior Nares samples collected directly into 750uL of Tris-EDTA (TE). All nasal swabs collected in this manner will be received in the UCLA COVID19 testing lab in 650 Charles E Young Dr South, CHS Building for direct accessioning and processing.

1) Nasal Swabs collected into pre filled tubes with 750uL TE are stored and transported at 4°C.
2) Tubes are placed into a 96 well rack and placed in a 95°C circulating water bath for 30 minutes.
3) Samples are spun in 4°C centrifuge for 20 seconds at 200xg to collect nasal swab sample at bottom of tube.
4) Rack is placed in the automated decapper (Brooks Intellecap or ThermoFisher Capit-All) and the 96 wells are uncapped.
5) Using the Benchsmart 20, we pipet 2.5 uL of Nuclease Free water, pipet an airgap of 2.5 uL, and then pipet 2.5uL of Extraction Free Nasal Swab. The total liquid volume of 5uL is dispensed into a designated quadrant of a 384-well plate pre-loaded with primer and mastermix.
6) After each quadrant is dispensed into the 384-well plate, the 384 well plate is placed into the thermocycler and run with the RT-PCR reaction described below.

#### 3.6. RT-PCR

##### 3.6.1. *Make Master Mix:*

(This calculation is for a single 384 well plates). This process is the same regardless of the sample type above (purified RNA from nasal swab, extraction-free nasal swab, extraction-free saliva)

Reagents:

TaqPath™ 1-Step RT-qPCR Master Mix (Fisher Scientific, A15300)

NUCLEASE-FREE WATER (Thermofisher, AM9937)

S Standard Dilution (see section G)

Table 7. MasterMix calculation.

| **Mastermix Calculation** | | | |
| --- | --- | --- | --- |
| **per 384 well plate** |  | | |
|  | **RT-PCR mix:** | **uL or copies per reaction** | **Total** |
|  | 4x Mastermix | 5 uL | 1920 |
|  | Water | 8 uL | 3702 |
| Dilution 4 | S RNA Standard quant  250 copies *384 | 192,000 copies | Calculate from dilution |
| Sample | 5 | |  |
| indexed primers | 2 | |  |
| Total Volume | 20 | | |
| Total Mastermix per well | 13 | | |

Notes:

- Master mix is made in a clean hood that has been treated for 15 minutes with UV light and cleaned with 10% bleach solution.
- All pipettes and water used are also treated with UV light

##### 3.6.2. *Tracking sample plate (refer to sample set up sheet)*

1) Each plate will be designated a quadrant: 1, 2, 3 or 4 of the 384-well plate. Record the Barcode and quadrant for each 96-well plate or tube-rack of samples.

##### 3.6.3. *Putting Together PCR Plate*

1) Clean benchtop surface with 10% Bleach
2) RNAse inhibitor treatment of benchtop surface
3) Take out pre-stamped primer plates (see section D) from -20 freezer and thaw on ice
4) Spin plates down at 2000xg for 1 minute
5) Visually inspect to ensure there is primer in each well
6) Pipet 13 uL of master-mix into each well using the Rainin Benchsmart 20 or PrePCR Integra Viaflo 12.5uL
7) Add 5ul of sample as described above in sections D, E and F for purified RNA from nasal swabs, extraction free saliva or extraction-free nasal swabs.

##### 3.6.4. *Reverse Transcription and PCR*

1. Program the following into the 384 Well Veriti Thermocycler.

Table 8. PCR cycle*.*

| 1 | 55C | 10 min |
| --- | --- | --- |
| 2 | 95C | 1 min |
| 3 | 95C | 10 sec |
| 4 | 60C | 30 sec |
|  | Go to step 3, 40 cycles (purified samples) or 50 cycles (unpurified samples) |  |
|  | Hold at 12C |  |

2. Load plate into 384 well cycler
3. Press Start

#### 3.7.  POST-PCR PROCESSING

#### 3.7.1. *PCR products Combined and purified.*

1) Use Integra Viaflow, pipet 6 ul from each well of a 384 well plate into a sterile reservoir.
2) Repeat step one for each 384 well plate that will be combined into a sequencing reaction run. All samples will be combined into the same reservoir.
3) Slightly tilt the reservoir back and forth to mix.
4) Transfer the entire volume to a 15 mL conical tube and vortex thoroughly.
5) Transfer 100 ul to an eppendorf tube.

##### 3.7.2. *Bead Cleanup*

1) Add 50 ul of AmpureXP beads (0.5:1 ratio of beads: sample volume) to 100 ul volume of pooled PCR reaction. Vortex and let sit for 5 minutes at room temperature.
2) Use magnet to collect beads for 1 minute.
3) Transfer supernatant (~150 ul) to a new eppendorf tube.
4) Add 130 ul of AmpureXP beads to the 150 ul of supernatant. Vortex and let sit for 5 minutes at room temperature.
5) Use magnet to collect beads for 1 minute.
6) Remove liquid and wash beads twice with 500 ul of freshly made 70% EtOH.
7) Elute in 40 ul of qiagen EB buffer
8) Use a magnet to collect beads for 1 minute. Transfer 33 ul to new eppendorf tube.

##### 3.7.3. *Library Quantification and Quality Control using Traditional S RNA Standard*

*Note: follow either this protocol or the one listed in the next section depending on which type of S RNA standard you plan to use (either traditional or diversified)*

1) Make a 1:10 dilution of eluted library using Ultrapure Water.
2) Use High Sensitivity DNA Qubit to measure the concentration of the 1:10 dilution.
3) Use the following link to determine the concentration of the 1:10 dilution in nM -<https://support.illumina.com/bulletins/2016/11/converting-ngl-to-nm-when-calculating-dsdna-library-concentration-.html>. Use 195bp for size.
4) Based on the above calculation, make a 5 nM dilution from the 1:10 dilution.
5) Measure this 5nM dilution and a stock of Illumina PhiX using High Sensitivity DNA Qubit.
6) Use the link from step 3 to calculate the concentration in nM of the 5nM dilution and the Illumina PhiX stock.
7) Make a dilution of the PhiX stock to equal the nM concentration of the 5nM dilution. For example, if the“5nM” dilution was actually 4.39 nM, dilute PhiX down to 4.39 nM.
8) Combine 14 ul of the “5 nM” dilution with 5ul of the dilution of PhiX. This results in a “5nM” library (4.39 nM in the example) that is 30% PhiX and 70% the library of interest.
9) Run the above 5nM library on an Agilent Technologies D1000 High Sensitivity Screentape.

##### 3.7.4. *Library Quantification and Quality Control using Diversified S RNA Standard*

*Note: follow either this protocol or the one listed in the next section depending on which type of S RNA standard you plan to use (either traditional or diversified)*

1) Make a 1:10 dilution of eluted library using Ultrapure Water.
2) Use High Sensitivity DNA Qubit to measure the concentration of the 1:10 dilution.
3) Use the following link to determine the concentration of the 1:10 dilution in nM -<https://support.illumina.com/bulletins/2016/11/converting-ngl-to-nm-when-calculating-dsdna-library-concentration-.html>. Use 195bp for size.
4) Based on the above calculation, make a 5 nM dilution from the 1:10 dilution.
5) Measure the concentration of the 5nM dilution using the High Sensitivity DNA Qubit
6) Use the link above to calculate the concentration in nM of the 5nM dilution. ***Note: with the diversified S RNA standard, do NOT quantify and add PhiX to the 5nM dilution. Proceed to the next section.***

#### 3.8. SEQUENCING PRIMER MIXES AND LOADING

#### 3.8.1.  *MiniSeq Primer Mixes*

a. Add 80 ul of water, 10 ul of S read 1 primer (100 uM stock), and 10 ul of RPP3 read 1 primer (100 uM stock) to an eppendorf tube labeled "Read 1 primer mix". Final concentration will be 20 uM of primers (10 uM of each read 1 primer).
b. Add 80 ul of water, 10 ul of S i7 primer (100 uM stock), and 10 ul of RPP3 i7 primer (100 uM stock) to an eppendorf tube labeled "i7 primer mix". Final concentration will be 20 uM of primers (10 uM of each i7 primer).
   3.8.2. ***MiniSeq Sequencing***

a. Load 24.5uL of the read 1 primer mix into reservoir 24. Mix.
b. Load 26uL of the i7 primer mix into reservoir 28. Mix.
c. Load 26uL of the i5 primer mix into reservoir 28. Mix.
d. Load 500uL 1.5pM library into reservoir 16.

##### 3.8.3. *NextSeq Primer Mixes*

a. Add 80 ul of water, 10 ul of S read 1 primer (100 uM stock), and 10 ul of RPP3 read 1 primer (100 uM stock) to an eppendorf tube labeled "Read 1 primer mix". Final concentration will be 20 uM of primers (10 uM of each read 1 primer).
b. In addition to the read 1 primer and i7 primer mix above, the NextSeq requires an i5 primer mix. To make this, add 80 ul of water, 10 ul of S i5 primer (100 uM stock), and 10 ul of RPP3 i5 primer (100 uM) stock to an eppendorf tube labeled “i5 primer mix”. Final concentration will be 20 uM of primers (10 uM of each i5 primer).

#### 3.8.4. *Next Seq Sequencing*

a. Load 52uL of the read 1 primer mix into reservoir 20. Mix.
b. Load 85uL of the i7 primer mix into reservoir 22. Mix.
c. Load 85uL of the i5 primer mix into reservoir 22. Mix.
d. Load 1300uL 1.25	pM library into reservoir 10.

### 4. **DATA ANALYSIS**

Illumina BCL files are downloaded and converted into FASTQ sequencing files using Illumina’s bcl2fastq software. Each amplicon sequence consists of a set of three individual reads: one 26 base pair read (read1) that identifies the amplicon (S, S standard, or RPP30) and two 10 base pair index reads (index1 and index2) that together uniquely identify the sample. Sequences are assigned to samples using the two index reads and the sum of the reads for each amplicon in each sample is obtained. Decisions about whether the sample passed QC and whether SAR-CoV-2 was detected in a sample are based on the count of sequences observed for each amplicon within each sample and explained in detail below.

##### 4.1. *MiSeq Control Software:*

The MiSeq Control Software (Illumina Inc., ‘For Research Use Only’) controls the flow cell

stage, temperature and fluidics system. It also captures images of clusters, generating image analysis, base calling, and base call quality data.

##### *4.2. Real Time Analysis Software*:

Primary analysis is performed by the Real Time Analysis (RTA) software (Illumina Inc., ‘For

Research Use Only’) and consists of base calling of each cluster at each cycle. In addition to base

calling, RTA assigns an analytical quality score (Q-score) to each base call. Calculations of

Q-scores are based on the ratio of the signal intensity of the highest base in a given cluster during a given cycle to the signal intensity of the three other bases. The quality score Q is calculated as -10 log10 P, where P is the probability that base call is incorrect. A minimum of 80% of basecalls must meet the Q30 threshold to proceed to data analysis. If these criteria are not met this could be due to a lack of sequence diversity (insufficient PhiX concentration), a technical problem in library construction, a faulty flow cell or sequencing instrument failure. If fewer than 80% of basecalls meet the Q30 threshold the entire run is discarded.

##### *4.3. bcl2fastq Conversion Software:*

The bcl2fastq conversion software (Illumina Inc., ‘For Research Use Only’) is used to process

BCL (base call log) files generated by the MiSeq instrument and convert time into FASTQ files. FASTQ is a standard text-based file format that will store the nucleotide sequences and base quality scores for each read sequenced from a sample. Three FASTQ files are generated, one corresponding to 26 base pairs of sequence within each amplicon (read1) and two 10 base pair index sequences (index1 and index2) that together uniquely identify each sample.

##### *4.4. UCLA Sample Demultiplexing and Amplicon Counting Software*:

Read1 is matched to one of the three expected amplicons allowing for the possibility of a single nucleotide error in the amplicon sequence. The set of three reads is discarded if read1 has a hamming distance greater than 1 from the expected amplicons. Samples are demultiplexed using the two index reads. Demultiplexing means assigning the sequences to the sample from which they originated. Observed index reads are matched to the expected index sequences allowing for the possibility a single nucleotide error in one or both of the index sequences. The set of three reads are discarded if both index1 and index2 have hamming distances greater than 1 from the expected index sequences. The sum of reads for each amplicon and each sample is calculated.

A fully automated R package that runs our Amplicon Counting Software and generates quality-control reports and interprets results for patient samples can be found at: <https://github.com/joshsbloom/swabseqr>

The development version of scripts for the initial implementation of our analysis pipeline and Amplicon Counting Software can be found at <https://github.com/joshsbloom/swabseq>

### 5. **RESULT INTERPRETATION FOR PURIFIED SAMPLES**

We require that 10 reads are detected for RPP30 for each sample. This serves as a control that sample collection took place properly and contains a human specimen. If fewer than 10 reads are detected for RPP30 the results are considered inconclusive.

We require that the sum of S and S synthetic standard reads exceeds 2,000 reads or the results are considered inconclusive. The S synthetic standard is added to the master mix, is present in every well and every sample in our assay, and even if no virus is present, if the primers and the assay are working properly the S synthetic standard will amplify and be sequenced. We have observed that samples with very high viral concentrations will result in high S reads and low S synthetic standard reads, and samples with low viral concentrations will result in low S reads and high S synthetic standard reads. In both cases the sum of S and S standard should be high in any sample regardless of the presence of Sars-CoV-2. This follows from the fact that the same S primers have equal preference for the S and S synthetic standard and amplify both with equal efficiency.

Assuming a sample as greater than 10 RPP30 reads and that the sum of S and S synthetic standard reads exceeds 2,000, we determine if SARS-CoV-2 is detected in a sample by seeing if the ratio of S to S standard exceeds 0.003. (We note that we add 1 count to both S and S standard before calculating this ratio to facilitate plotting the results on a logarithmic scale.) If the ratio is greater than 0.003 we concluded that Sars-CoV-2 is detected for that sample and if it is less than or equal to 0.003 we conclude that Sars-CoV-2 is not detected.

Table 9. Result interpretation for purified samples

| **Well-controls** | |  | **Results** | | | |
| --- | --- | --- | --- | --- | --- | --- |
| **Total S + S Standard** | **RPP30 read count** |  | **S/S Standard ratio** | **Result** | **Interpretation** | **Action** |
| >2000 reads | >10 |  | > 0.003 | SARS- CoV-2  Detected | Positive for SARS-CoV-2 for the Sample ID. | Report results to physician, patient, and appropriate public health authorities. |
| >2000 reads | >10 |  | < 0.003 | SARS- CoV-2  Not Detected | Negative for SARS-CoV-2 for the Sample ID. | Report results to physician, patient, and appropriate public health authorities. |
| <2000 reads | >10 |  | - | Inconclusive | Invalid for the Sample ID. | Quality control for the Sample ID is FAIL. Repeat sample or Recollect sample |
| >2000 reads | < 10 |  | - | Inconclusive | Invalid for the Sample ID. | Quality control for the Sample ID is FAIL. Repeat sample or Recollect sample |
| <2000 reads | < 10 |  | - | Inconclusive | Invalid for the Sample ID. | Quality control for the Sample ID is FAIL. Repeat sample or Recollect sample. |

### 6. **RESULT INTERPRETATION FOR EXTRACTION-FREE SALIVA AND NASAL SWAB**

We require that the sum of S and S synthetic standard reads exceeds 500 reads, or the results are considered inconclusive. With inhibitory lysates we have observed that this lower total is acceptable for maintaining sensitivity and specificity while limiting the number of tests that are considered inconclusive. If the sum of S and S synthetic standard reads exceeds 500, we determine if SARS-CoV-2 is detected in a sample by seeing if the ratio of S to S standard exceeds 0.0	5. (We note that we add 1 count to both S and S standard before calculating this ratio to facilitate plotting the results on a logarithmic scale.) If the ratio is greater than 0.05	we concluded that Sars-CoV-2 is detected for that sample and if it is less than or equal to 0.05	we conclude that Sars-CoV-2 is not detected as long as 10 reads are detected for RPP30 for that sample. This serves as a control that sample collection took place properly and contains a human specimen. If fewer than 10 reads are detected for RPP30 and the ratio of S to S standard is less than or equal to 0.05	the results are considered inconclusive. We have modified this criteria such that only samples without Sars-CoV-2 signal are considered inconclusive if RPP30 reads are fewer than 10. This ensures that we do not miss Sars-Co	V-2 positive samples that may have fewer RPP30 reads.

Table 10. Result interpretation for extraction-free saliva and nasal swab samples

| **Well-controls** | |  | **Results** | | | |
| --- | --- | --- | --- | --- | --- | --- |
| **Total S + S Standard** | **RPP30 read count** |  | **S/S Standard ratio** | **Result** | **Interpretation** | **Action** |
| >500 reads | >0 |  | > 0.05 | SARS- CoV-2  Detected | Positive for SARS-CoV-2 for the Sample ID. | Report results to physician, patient, and appropriate public health authorities. |
| >500 reads | >10 |  | < 0.05 | SARS- CoV-2  Not Detected | Presumptive Negative for SARS-CoV-2 for the Sample ID. | Report results to physician, patient, and appropriate public health authorities. |
| <500 reads | >0 |  | - | Inconclusive | Invalid for the Sample ID. | Quality control for the Sample ID is FAIL. Repeat sample or Recollect sample |
| >500 reads | < 10 |  | <.05 | Inconclusive | Invalid for the Sample ID. | Quality control for the Sample ID is FAIL. Repeat sample or Recollect sample |
